# Association of childhood vaccination with family planning, healthcare access, and women education: analysis of Nepal, Senegal and Zambia

**DOI:** 10.1101/2022.11.26.22282789

**Authors:** Francisco Castillo-Zunino, Kyra A Hester, Pinar Keskinocak, Dima Nazzal, Matthew C Freeman

## Abstract

**Background:** Childhood vaccination, family planning, healthcare access, and women’s empowerment are goals targeted by the Sustainable Development Goals (SDG). Barriers to healthcare access impede vaccination, and tackling goals holistically could create larger gains than siloed efforts. We studied Nepal, Senegal, and Zambia to test the association between childhood vaccinations and other SDG indicators to identify clustered deprivations. We quantified how children with few – or no – vaccines and their mothers were vulnerable in other SDG areas.

**Methods:** We analyzed Demographic and Health Surveys (DHS) from Nepal, Senegal, and Zambia. Through ordinal logistic regressions, controlling for household/mother’s characteristics, we identified strong predictors of the number of vaccine doses one-year-old children received. Through bootstrapping and optimal propensity scores matching, we compared children with no or few childhood doses (0-2 doses in early 2000s, or 0-4 in late 2010s) to children who received eight doses (DTP1-3, MVC1, Pol1-3, and BCG vaccines).

**Findings:** Mothers of children who received eight doses were 14-30% more likely than mothers of children with few or no doses to have accessed a health facility in the last year (95% CIs were 16-44% in Nepal 2001, -5% to 33% Nepal 2016, 3-26% Senegal 2005, 1-31% Senegal 2019, 9-38% Zambia 2001-02, 7-36% Zambia 2018), knew on average 0.7-1.5 more contraceptive methods (0.9-2.0 Nepal 2005, 0.1-1.5 Nepal 2016, 0.6-1.7 Senegal 2005, 0.2-1.7 Senegal 2019, 0.1-1.4 Zambia 2001-02, 0.5-1.4 Zambia 2018), and had 10-22% higher literacy rates (12-32% Nepal 2001, -7% to 36% Nepal 2016, 10-26% Senegal 2005, -3 to 22% Senegal 2019, -4% to 28% Zambia 2001-02, 5-36% Zambia 2018).

**Interpretation:** Children with few or no vaccine doses and their mothers were behind in access to family planning, healthcare, and education compared to fully vaccinated children and their mothers. Such differences can further impede immunizations; therefore, integrated education and health services are needed to improve vaccination outcomes.

**SUMMARY**

What is already known about this subject?

- Effective integrated health services have been implemented to simultaneously improve multiple health outcomes and other Sustainable Development Goals (SDG).
- Childhood vaccinations should be integrated with other initiatives, rather than keeping activities siloed, to further achieve SDG goals.

What are the new findings?

- Mothers and their children who received few – or no – vaccines in Nepal, Senegal, and Zambia share multiple vulnerabilities related to access to family planning, access to health facilities, and health/women education.

What are the recommendations for policy and practice?

- There is an opportunity to integrate vaccination efforts with other healthcare services and women’s education initiatives to increase performance outcomes of vaccination coverage and other SDG goals.

## Introduction

The global coverage of the third dose of the diphtheria-tetanus-pertussis vaccine (DTP3s) dropped from 86% in 2019 to 81% in 2021, having the highest number of children not receiving recommended immunization since 2009 – further deviating from the United Nation’s Sustainable Development Goals (SDGs) and Immunization Agenda 2030 [1]. Low- and middle-income countries are disproportionately affected by inequities in healthcare access [2,3]. Intersectional inequalities exist between health outcomes and other vulnerabilities [4]; for example, education plays a key role in improving economic and health outcomes [5]. To improve health outcomes, such as decreasing childhood mortality, it is necessary to understand and quantify clustered deprivations.

Efforts to tackle vulnerabilities can focus on a single issue or follow an integrated strategy by which multiple issues are addressed simultaneously. Evidence suggests that improving maternal education and women’s empowerment [6, 7, 8] or integrating family planning services with routine childhood immunization generates successful outcomes [9, 10]. To achieve equitable healthcare access, improvements need to focus on hard-to-reach children and women affected by clustered deprivations [11], ideally through integrated approaches that target SDGs more efficiently than current siloed efforts.

SDGs provide 17 goals and 169 targets with the overarching aim of “a world free from poverty, hunger and disease.” Goal 3 is to improve health and general well-being and contains the target of reducing child mortality – wherein routine childhood vaccination has been a highly effective intervention [12,13,14]. SDG5 aims to achieve gender equality, where women’s empowerment has been relevant to improve childhood vaccinations in low- and middle-income countries [15].

The objective of this study was to quantify the extent to which childhood immunization was associated with other health and development indicators. We studied factors related to family planning (education and access), reproductive and child healthcare access, and women’s education and empowerment. By quantifying the differences between children who received few vaccine doses compared to fully vaccinated children, we identified clustered deprivations where integrated approaches have a potential to improve multiple vulnerabilities simultaneously. We used data from Nepal, Senegal, and Zambia – three low- and lower-middle-income countries that achieved substantial growth in vaccination coverage rate in the last twenty years [16].

Studies that analyze the association between vaccination and other healthcare outcomes typically use single vaccines – such as DTP1 and DTP3 – as proxies for the immunization system; wherein DTP1 is a proxy for initial access to the system, and DTP3 is proxy for continued engagement with the system. In the more rigorous cases, studies analyze additional childhood vaccines by repeating the statistical analysis for each vaccine type independently [7]. Using the coverage rate for a single vaccine dose, such as DTP1 or DTP3, does not lead to accurate estimates for the rate of children who received none or all 8 routine childhood immunizations.

To our knowledge, our study was the first to analyze the coverage rates for childhood vaccines, considering the 8 routine immunization doses in an ordinal manner from children that received 0 to all 8 vaccines. This statistical consideration is relevant, as efforts needed for children to get an additional vaccine is not the same when they have 0 vaccines compared to when they have 7. The ordinal logic enabled us to analyze the entire vaccination system more granularly to identify factors that impact childhood immunization coverage.

## Methods

### Patient and public involvement

Patients were not involved.

### Overview

We used ordinal logistic regression models (OLR) [17, 18] to test for strong associations between vaccine doses and other health indicators in Nepal, Senegal, and Zambia. We employed bootstrapping [19] combined with optimal propensity scores matching [20,21,22] to compare children with few or no vaccine doses (we consider few as receiving 0-2 doses in early 2000s, or 0-4 doses in late 2010s) with similar children who were fully vaccinated. We consider a child to be fully vaccinated as having received the following 8 vaccine doses: first to third doses of diphtheria-tetanus-pertussis vaccine (DTP1 to DTP3), BCG vaccine, first dose of measles-containing vaccine (MVC1), and first to third doses of the polio vaccine (Pol1 to Pol3). This paper focuses on the following areas of interest: family planning education/access (A1), reproductive & child healthcare access (A2), and woman education & empowerment (A3).

This study was part of the vaccine exemplars study, which assessed how and why some countries achieved high growth and sustained coverage of early childhood vaccination [23]. Using a case study positive deviant approach, we identified three such countries based on available country-level data on early childhood vaccination (Nepal, Senegal, and Zambia) [24,25,26]. Additional details of the selection of these countries can be found elsewhere [27].

### DHS data filtering & year selection

The Demographic and Health Surveys Program (DHS) has collected representative household-mother-child level data on population characteristics, childhood vaccinations, and data related to areas A1, A2, and A3 over the years since 1984 [20]. We used child, mother, household, and wealth data (named respectively by the DHS Program: BR, IR, HR, and WI databases) as main data sources for Nepal, Senegal, and Zambia. The data can be accessed upon approval from the DHS Program [28]. Based on the DHS Program recommendations to study childhood vaccinations [29], we worked with a subset of surveyed children who were alive and one-year-old; by filtering variables *b5* = ‘yes’ and *b8* = 1.

We considered DHS surveys from 2000 to 2020 for analysis, selecting the earliest and latest surveys available for each country (see Figure 1). Nepal conducted DHS surveys in 2001, ‘06, ‘11, and ‘16; we selected years 2001 (72% DHS DTP3 coverage) and 2016 (86%) for our analysis. Senegal surveyed mothers in 2005, ‘10-11, and later conducted continuous surveys through 2012-19; we selected years 2005 (78%) and 2019 (92%). Zambia had DHS surveys in 2001-02, ‘07, ‘13-14, and ‘18; we selected 2001-02 (80%) and 2018 (92%). We started our analysis from year 2000 to align with the creation of Gavi, launch of the Millennium Development Goals [30], and because older DHS surveys differed drastically with more current surveys.

**Figure 1:**
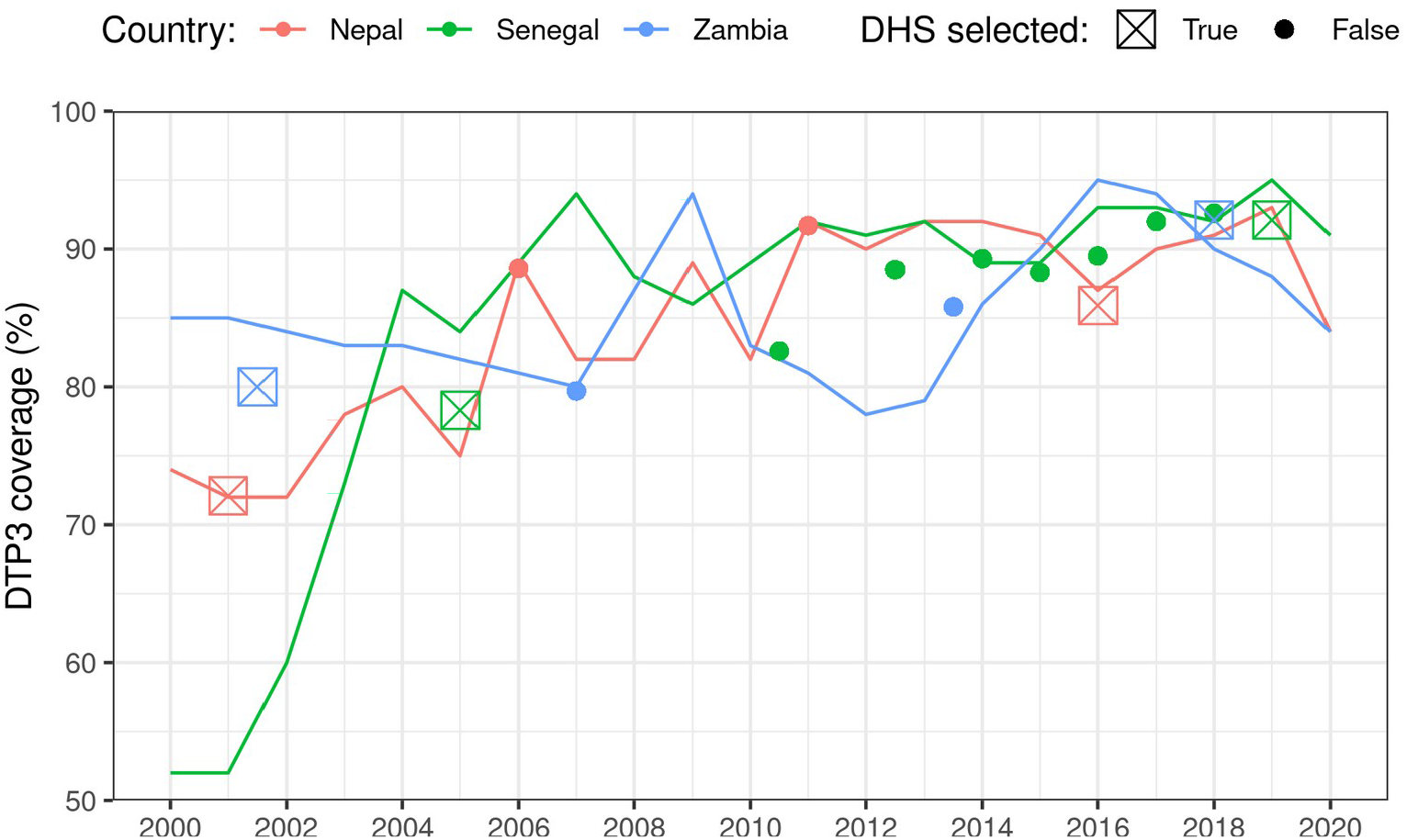
Selected DHS surveys and DTP3 coverage of Nepal, Senegal, and Zambia, years 2000-2020 Data sources: Various DHS surveys [28] and WHO/UNICEF estimates of national infant immunization coverage [16]. Points represent DTP3 coverage estimates from DHS, squares highlight the DHS-years selected for analysis, and lines show the WHO/UNICEF estimates of DTP3 coverage.

### Grouping children by number of vaccine doses

Immunization studies typically use the coverage rates of proxy immunizations such as DTP1 and DTP3 to estimate the rates of children who started receiving and completed the recommended routine immunization, respectively; this is common practice due to the number and timing of the doses of the vaccines included in the recommended childhood vaccination schedules [16]. While this approach simplifies data requirements and analysis, it lacks granularity regarding how many children received no or all recommended immunizations, or any number (receiving a subset of recommended immunizations) in between, limiting statistical analysis and our understanding of factors that might influence vaccination coverage. Utilizing DHS data, we analyzed DTP1-3, BCG, MVC1, and Pol1-3 due to their inclusion in Nepal, Senegal, and Zambia’s childhood immunization programs; these vaccines were introduced in each country in 1977 as part of the Expanded Programme on Immunization [31]. By analyzing the different number of vaccine doses received by children, we can better understand their progress towards being fully vaccinated and the factors that impact this progress.

We categorized all one-year-old children into one of three groups: few or no vaccine doses (LV, as in low vaccination level), all 8 vaccine doses (8V), and anything in between (MV, as in middle vaccination level). Group LV was defined differently depending on the DHS-year (0-2 or 0-4 doses), since there were very few samples for 0-2 vaccine doses after 2010; particularly in Nepal 2016 with only 19 samples. For DHS datasets before 2010, we defined LV as children with 0 to 2 doses; MV corresponds to 3-7 doses. For DHS surveys after 2010, we defined LV as 0-4 vaccine doses; MV as 5-7 doses. Table 1 shows the number of samples per number of vaccine doses; note group LV had 4 to 7% of the one-year-old children sampled on each DHS dataset^2^.

**Table 1:** Sample size of one-year-old children per number of vaccine doses

| DHS dataset |  | Number of one-year-old children sampled per 0 to 8 vaccine doses |  |  |  |  |  |  |  |  |  | Percentage of one-year-old children sampled per vaccine group |  |  |
| --- | --- | --- | --- | --- | --- | --- | --- | --- | --- | --- | --- | --- | --- | --- |
| Country | Year | 0 | 1 | 2 | 3 | 4 | 5 | 6 | 7 | 8 | Total | LV | MV | 8V |
| Nepal | 2001<sup>†</sup> | 10 | 5 | 75 | 112 | 28 | 54 | 58 | 101 | 856 | 1 299 | 6.9 | 27.2 | 65.9 |
|  | 2016* | 10 | 6 | 3 | 8 | 15 | 31 | 45 | 96 | 811 | 1 025 | 4.1 | 16.8 | 79.1 |
| Senegal | 2005<sup>†</sup> | 88 | 27 | 29 | 62 | 38 | 161 | 123 | 349 | 1 263 | 2 140 | 6.7 | 34.3 | 59.0 |
|  | 2019* | 38 | 7 | 4 | 18 | 10 | 30 | 20 | 154 | 902 | 1 183 | 6.5 | 17.2 | 76.2 |
| Zambia | 2001-02<sup>†</sup> | 44 | 8 | 17 | 32 | 24 | 61 | 81 | 147 | 915 | 1 329 | 5.2 | 26.0 | 68.8 |
|  | 2018* | 28 | 4 | 11 | 24 | 20 | 41 | 68 | 285 | 1 447 | 1 928 | 4.5 | 20.4 | 75.1 |

*Data sources: Various DHS surveys [28].*
<sup>†</sup> denotes DHS surveys where LV consists of one-year-old children with 0-2 vaccines and MV of 3-7 vaccine doses.
\* denotes DHS surveys where LV consists of one-year-old children with 0-4 vaccines and MV of 5-7 vaccine doses.

### Variable selection & statistical significance

We selected variables from DHS data; either for use as control variables for regression and propensity score matching, or for analysis to determine key outcomes and to quantify the vulnerabilities shared by LV children. Table 1 in Supplemental Materials shows which DHS variables were selected, their DHS categories and sub-categories [29], and if they were used as control variables. We only selected variables available and comparable in all selected DHS surveys for Nepal, Senegal, and Zambia. Table 2 in Supplemental Materials shows in detail how each DHS variable was processed and interpreted.

#### Control variables

We selected a total of 14 control variables related to household and mother’s characteristics. The control variables related to household characteristics were: (1) number of residents, (2) have improved source of drinking water, (3) have improved sanitation facility, (4) not sharing toilet with other households, (5) has radio, (6) has TV, (7) has electricity, (8) has bicycle, (9) urban or rural area, and (10) wealth index factor. Control variables related to mother’s characteristics were: (11) age, (12) has partner, (13) works or worked in past year, and (14) number of children ever born. These control variables limit the influence of living standards on analysis results.

We calculated the Variance Inflation Factor (VIF) among the 14 control variables, and identified no multicollinearity issues (all values <10). Table 3 in Supplemental Materials shows the VIF for each control variable, country, and year combination.

#### Key outcomes

We selected 38 variables of interest from the following DHS categories: family planning & fertility (related to area A1), reproductive & child health access (A2), women’s empowerment, and mother’s education (A3); see Table 1 in Supplemental Materials for full list of variables. We tested each indicator to find strong associations with immunizations through ordinal logistic regressions (OLR) [17, 18]. We used the groups LV, MV, and 8V as ordinal categories, the tested variable as independent variable (each tested separately), while controlling for all 14 control variables on every one of the six selected DHS datasets. The survey samples used to fit the OLRs were weighted according to the survey’s sample weights, as defined by DHS.

The 38 variables were also used to calculate the difference between 8V and LV children to identify clustered deprivations among LV children.

### Comparing children based on number of vaccine doses

#### Matching children

We matched 8V and MV children with LV children to account for the differences in living standards; those with few or no vaccine doses generally have worse living standards than children who received more vaccines [32]. Without matching, bias would overestimate the differences between our groups of vaccinated children; matching was based on the above 14 control variables.

For every vaccination group – LV, MV, and 8V – we calculated the average values for each variable. We resampled through a weighted bootstrap method (1000 replications) to obtain 95% CIs of the average estimations [19].

Next, we used the same weighted bootstrap method, but we used an optimal matching through propensity scores for each replication [20,21]; matching based on the selected control variables. Propensity score matching allowed us to obtain an unbiased estimation of the effect of childhood vaccinations on key outcomes – potentially identifying opportunities for implementing integrated policy efforts. The LV group was used as the baseline, so matched samples were obtained from MV and 8V children that most closely resemble children in LV. This allowed us to recompute the average values for each vaccine group while removing potential biases from any differences in living conditions.

#### Comparing groups LV and 8V

To know if children with few vaccine doses (LV) share common vulnerabilities different from fully-vaccinated children (8V), we compared the average differences between groups LV and 8V. Any important differences could be part of an integrated policy strategy to tackle multiple vulnerabilities – including low immunization coverage – on children with few or no vaccines, and vice-versa.

We compared the difference between the average values of all SDG variables and of LV and 8V by calculating the CI of the difference. This process identified variables where the differences between children in LV and 8V were statistically significant (95% CI). We calculated the averages and differences through 1000 bootstrap replications, with and without matching children in 8V and LV.

## Results

### Control variables & LV characteristics

Mothers of children in LV compared to those in 8V had 0.6-1.6 more children on average (range given by average differences of all six DHS surveys). Children in LV, compared to those in 8V, had a wealth index 3-24 percentiles lower (suggesting worse economic conditions for the LV group) and were 5-14% more likely to be located in rural areas. LV household were 8-27% less likely to own a TV.

Supplemental Materials: Table 4 shows the CI of the coefficients of the 14 control variables for each DHS dataset, and Table 5 shows more details on the 8V and LV comparison of control variables.

### Identifying variables strongly associated with vaccine doses

Table 6 in Supplemental Materials shows a summary of the coefficients obtained by each of the 38 variables on each DHS dataset, with their respective 90% and 95% CIs. Table 7 in Supplemental materials shows a summary of the results.

Combining the OLR tests with the 8V-LV groups statistical differences, we identified which variables related to family planning (A1), reproductive & child healthcare access (A2), and woman empowerment & education (A3) had strong associations with vaccinations. We present results for the most relevant variables.

### A1: Family planning

Our analysis revealed strong, positive associations between vaccine doses and family planning knowledge and access. Figure 2 shows more details on both variables described. Mothers of children in 8V compared to LV:

**Figure 2:**
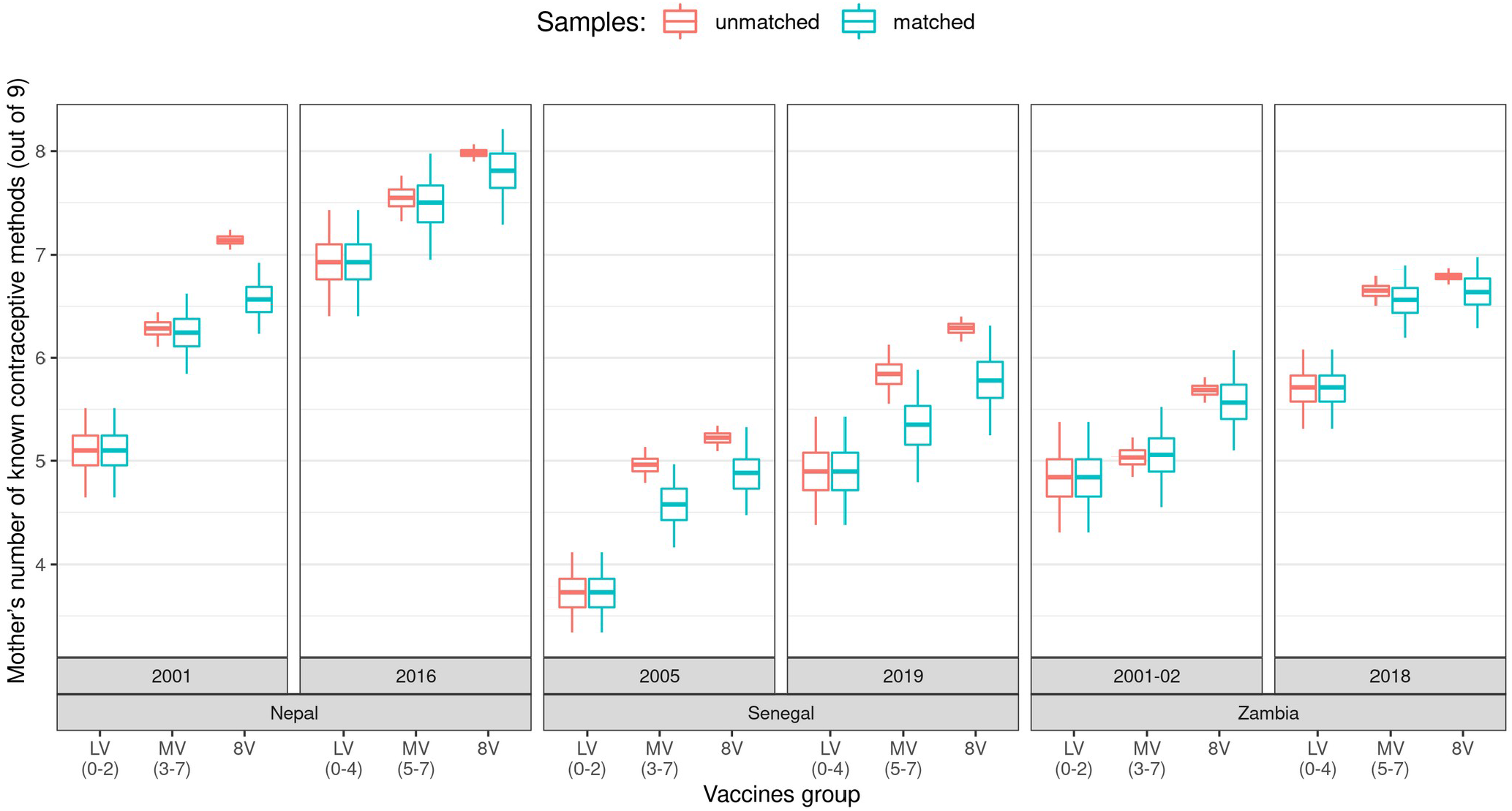
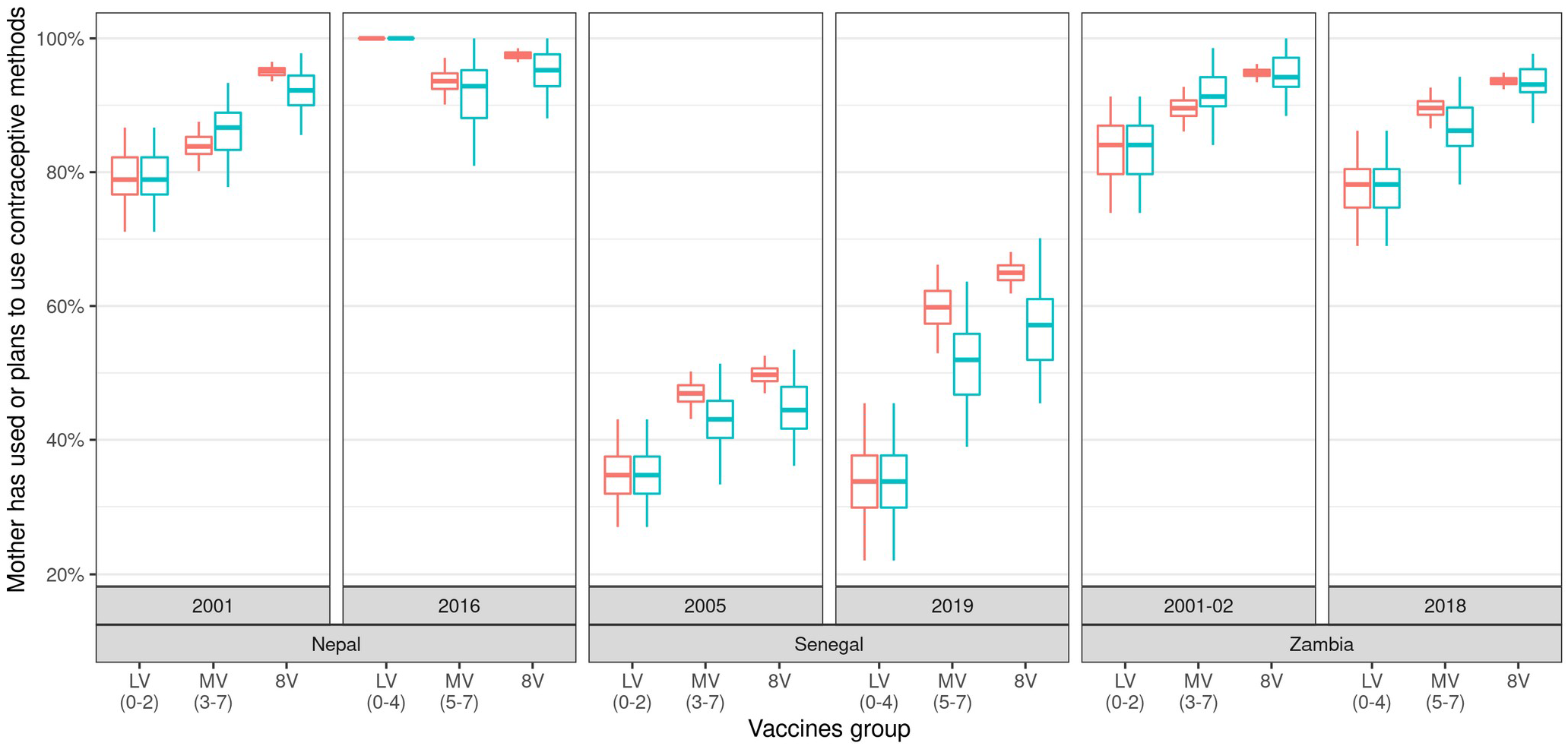
Number of contraceptives known by mother and if mother has used or plans to use contraceptive methods, by country, year, and number of vaccines groups Data sources: Various DHS surveys [28]. Each box-plot represents the average value for a specific country, year, and vaccine group either for matched or unmatched average calculations. Box-plots denote percentiles 25, 50, and 75, and their whiskers represent percentiles 2.5 and 97.5 (95% CI range). Red/left box-plots correspond to averages calculated through 1000 replications of weighted bootstraps, while cyan/right box-plots correspond to averages calculated through 1000 replications of weighted bootstraps while controlling for household/mother’s characteristics.

- knew on average 0.7-1.5 (range given by all six DHS averages) more contraceptive methods (95% CI ranges: 0.9-2.0 in Nepal 2005, 0.1-1.5 in Nepal 2016, 0.6-1.7 in Senegal 2005, 0.1-1.7 in Senegal 2019, 0.1-1.4 in Zambia 2001-02, and 0.5-1.4 in Zambia 2018)
- were 10-23%^3^ more likely to have used or plan to use contraceptives (95% CIs: 3-23% in Nepal 2005, -12% to 0% in Nepal 2016, -1% to 21% in Senegal 2005, 6-40% in Senegal 2019, 1-22% in Zambia 2001-02, and 6-26% in Zambia 2018).

Other family planning variables were also positively (but not strongly) associated with vaccine doses. Mothers of children in 8V compared to LV were:

- 3-30% more likely to use “modern contraceptive method”
- 0-26% more likely to have “heard about heard about family planning in the radio”
- 3-21% more likely to be “told side-effects when getting contraceptives”
- 2-20% more likely to know “her ovulatory cycle”

(Ranges given by all six DHS averages, refer to Table 7 in Supplemental materials to see the CI of each specific country and year.)

### A2: Reproductive & child healthcare access

A strong and positive association was found between vaccine coverage and mothers that accessed a health facility at least once during the last year prior to being surveyed, where 8V mothers were 17-30% more likely to have accessed health facilities than LV (range given by all six DHS averages); see Figure 3 for more details. The 95% CI for each DHS were: 16-44% in Nepal 2001, -5% to 33% in Nepal 2016, 3-26% in Senegal 2005, 1-31% in Senegal 2019, 9-38% in Zambia 2001-02, 7-36% in Zambia 2018.

**Figure 3:**
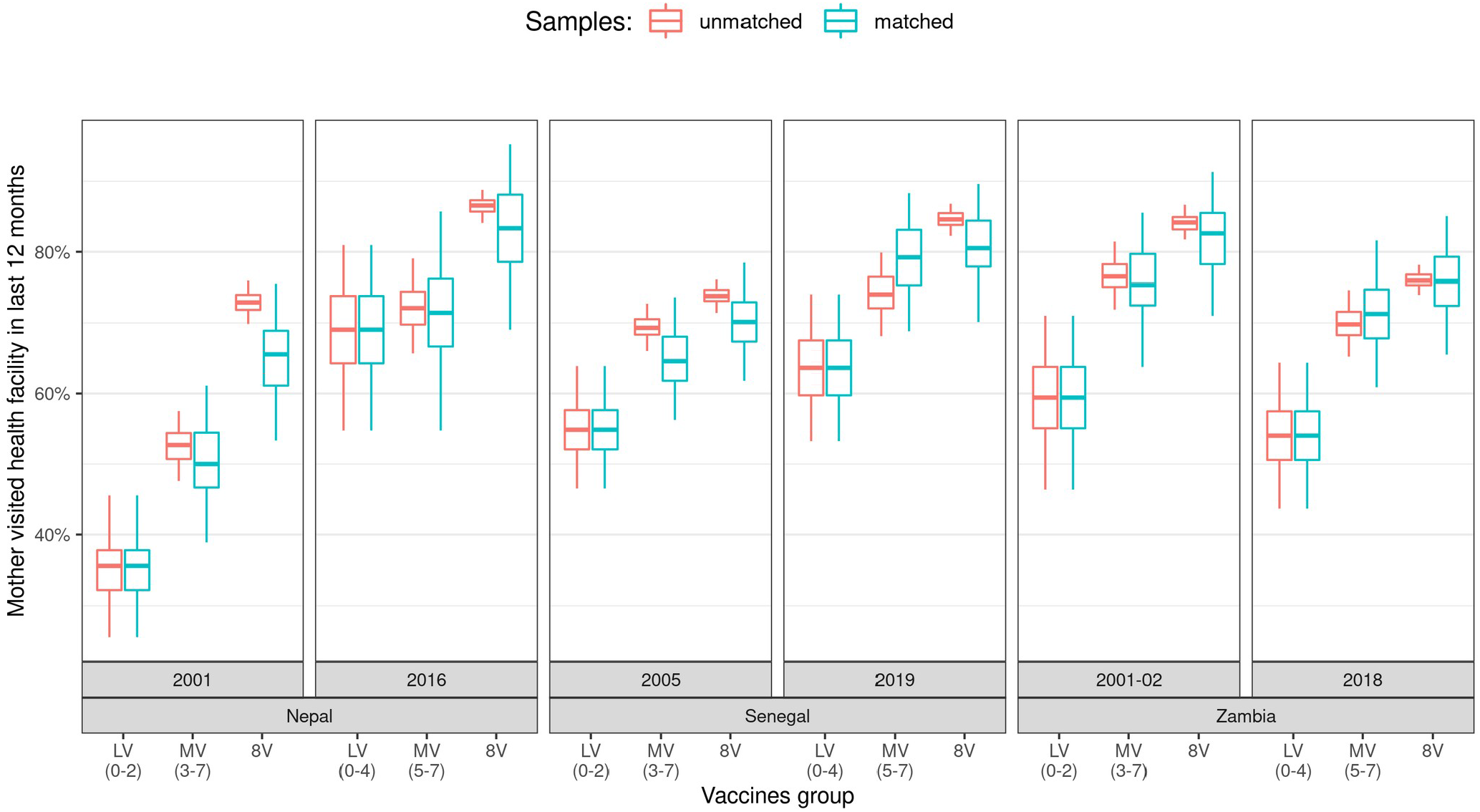
Percentages that visited a health facility during the last year, by country, year, and number of vaccines groups Data sources: Various DHS surveys [28]. Each box-plot represents the average value for a specific country, year, and vaccine group either for matched or unmatched average calculations. Box-plots denote percentiles 25, 50, and 75, and their whiskers represent percentiles 2.5 and 97.5 (95% CI range). Red/left box-plots correspond to averages calculated through 1000 replications of weighted bootstraps, while cyan/right box-plots correspond to averages calculated through 1000 replications of weighted bootstraps while controlling for household/mother’s characteristics.

Other reproductive & child healthcare access variables were also positively (but not strongly) associated with vaccine doses. Mothers of children in 8V compared to LV were:

- 6-39% more likely to “had at least 1 antenatal visit during pregnancy”
- 3-25% more likely to “had blood or urine sample taken during pregnancy”
- 2-21% more likely to “know Oral Rehydration Salts packets”

(Ranges given by all six DHS averages, please refer to Table 7 in Supplemental materials to see the CI of each specific country and year.)

### A3: Women’s education & empowerment

A positive association was found between vaccine doses and mother’s education, where mothers of children in 8V versus LV had:

- 10-22% higher literacy rates (95% CIs: 12-32% in Nepal 2001, -7% to 36% in Nepal 2016, 10-26% in Senegal 2005, -3% to 22% in Senegal 2019, -4% to 28% in Zambia 2001-02, 5-36% in Zambia 2018). Figure 4 shows the CI of the estimated mother literacy rates for each DHS.
- 0.7-1.5 more years of education (95% CIs: 0.2-1.3 in Nepal 2001, -0.6 to 2.1 in Nepal 2016, 0.6-1.7 in Senegal 2005, -0.3 to 1.6 in Senegal 2019, -0.2 to 1.8 in Zambia 2001-02, and 0.6-2.4 in Zambia 2018).

**Figure 4:**
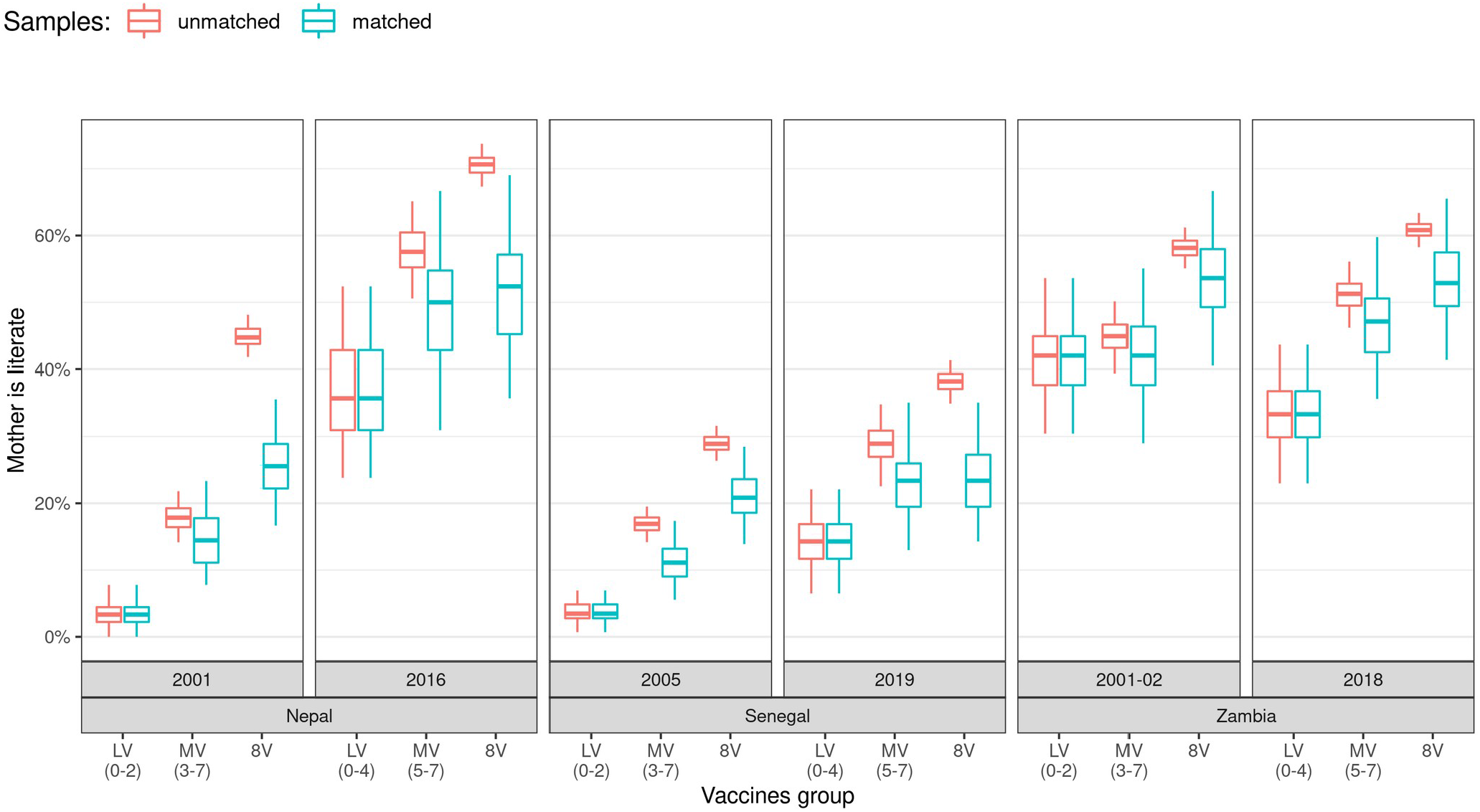
Mother literacy by country, year, and number of vaccines groups Data sources: Various DHS surveys [28]. Each box-plot represents the average value for a specific country, year, and vaccine group either for matched or unmatched average calculations. Box-plots denote percentiles 25, 50, and 75, and their whiskers represent percentiles 2.5 and 97.5 (95% CI range). Red/left box-plots correspond to averages calculated through 1000 replications of weighted bootstraps, while cyan/right box-plots correspond to averages calculated through 1000 replications of weighted bootstraps while controlling for household/mother’s characteristics.

There was no clear association between vaccine doses with domestic violence and mothers participating in decision making – decisions about child’s health, visiting relatives, making purchases, etc.

## Discussion

We conducted an analysis to identify factors that were strongly associated with routine childhood immunization coverage to determine if those factors were commonly shared between children who received few or no immunization doses. The analyses included data from three countries – Zambia, Nepal, and Senegal – that have relatively robust growth in early childhood vaccination rates relative to their peers.

Our three key findings revealed that the number of vaccine doses one-year-old children received was positively associated with: their mother’s knowledge and access to family planning services; access to health facilities and other reproductive and child health services; and their mother’s literacy and years of education. These associations persisted when comparing children who received all or few recommended immunizations (groups 8V and LV respectively) – even after matching 8V children to LV children, who overall came from worse economic situations, had fewer amenities (such as TV, radio, improved water), and who live more often in rural areas.

Controlling for mother and household characteristics allowed us to decrease biases that could overestimate the 8V-LV differences. Results involving domestic violence and mother’s decision making – as opposed to partners or someone else making decisions for them – were inconclusive and weakly associated with child vaccination doses received. These findings have policy implications, as stakeholders can prioritize funding and additional resources on the vulnerabilities that can be effectively siloed.

Mothers of children in LV versus 8V had lower access to family planning and other healthcare services. The findings of this study further highlight the existence of clustered deprivations in vaccination coverage and overall health and further highlight the importance of simultaneously addressing multiple SDGs while avoiding duplicate efforts [33]; for example, integrating supply chains is one strategy to improve health services [34]. Integrated efforts could potentially improve vaccine coverage by utilizing other healthcare services to provide mother with vaccination education, and vice-versa, where vaccination efforts can be used to educate mothers on other health services. For example, a study at a hospital in Pokhara, Nepal, identified that 14% of mothers (of children under 5 years old) that visited their facilities in 2015 were not aware of any family planning methods, and 19% were not familiar with the government’s childhood immunization schedule; a good example of how healthcare accessibility can be paired with mother health education to synergize with vaccine adoption and other health outcomes [35]. In Senegal, a study identified a strong and positive correlation between mother’s general education level and uptake of several vaccines [36].

Integrated efforts to address cluster deprivation can be executed successfully [9, 10, 37] and global guidelines are now shifting to recommend integrated care [38,39]. However, not all challenges can be addressed through integrated health services, and not all such implementations have been successful [40] – good practices must be followed to achieve success [41,42]. The strong associations found in this study – within three countries successful in vaccination coverage – give further evidence to consider integrated solutions to vaccine services.

## Conclusion

The results of this study show strong association between childhood immunization coverage and factors related to family planning services, reproductive & child health services, and woman’s education. Children who have received few or no vaccines (and their mothers) were also vulnerable in other areas; hence, the results highlight the importance of integrated policies and their implementation, addressing multiple issues. Nepal, Senegal, and Zambia are examples of low- and lower-middle-income countries with successful childhood immunization systems where governments and partner organizations have implemented solutions to improve vaccination coverage, while also focusing on healthcare access and women’s empowerment. Our results suggest that family planning efforts are more strongly associated with childhood immunization than other healthcare areas.

## Data Availability

Data are available upon request from DHS repositories.

https://dhsprogram.com/data/Using-Datasets-for-Analysis.cfm

## Acknowledgments

The authors would like to thank Zoe Sakas, Anna Rapp, Robert Bednarczyk, Gloria Ikilezi, Nancy Fullman, Nicoleta Serban, and Yajun Mei for feedback and discussion that helped with the content and exploration of this document. We appreciate the Demographic and Health Survey Program for allowing access to the data.

## Contributors

FCZ, in collaboration with PK and DN, did the analysis and wrote the first draft. KAH worked on improving the document. All authors contributed to the outline and revisions of the manuscript. MCF was the principal investigator of the Bill & Melinda Gates Foundation-funded research, and PK was one of the lead investigators.

## Funding

This work was supported by the Bill & Melinda Gates Foundation (OPP1195041). This research has also been supported in part by the William W. George endowment and the following benefactors at Georgia Tech: Andrea Laliberte, Joseph C. Mello, Richard E. & Charlene Zalesky, and Claudia & Paul Raines.

## Disclaimer

The funders played no role in data collection, analysis and formulation of results.

## Competing interests

None declared.

## Patient consent for publication

Not required.

## Provenance and peer review

Not commissioned; externally peer reviewed.

## Data availability statement

Data are available upon request from DHS repositories.

## Footnotes

2 We aimed to achieve that 5% of the children with fewer vaccines were in LV, but the 0-2 or 0-4 vaccine groups gave close percentages without having different vaccination-count ranges for each country-year combination

3 Range ignoring Nepal 2016, where LV and 8V were almost identical and above 90%.

## Notes

### Competing Interest Statement

The authors have declared no competing interest.

### Author Declarations

Demographic and Health Surveys (DHS) from Nepal, Senegal, and Zambia

