## Supplemental materials for "Association of childhood vaccination with family planning, healthcare access, and women education: analysis of Nepal, Senegal and Zambia"

#### A: Processing DHS variables

Table 1 shows all selected variables with information about their DHS categories and sub-categories, DHS database used, and if they were picked as control variables or not. Table 2 shows a summary of how each variable was computed based on DHS variables. We followed most suggestions given by the DHS program to calculate variables [1, 2]. Some minor changes were sometimes made, since DHS datasets are from different years/countries and variables are not always identically defined. Note we applied *log* –or *log(x+1)*– transformations to normalize some numerical variables in the regression models.

*Table 1: Variables selected from DHS data, sorted by DHS category*

| Sub-Category | DHS database | DHS variables used | Variables | Control? |
|---|---|---|---|---|
| **2) Population & Housing** | | | | |
| Household Composition | HR | hv012 | Household's number of residents | X |
| Household drinking water | HR | hv201 | Household has improved source of drinking water | X |
| Household Possessions | HR | hv207 | Household possesses radio | X |
| | HR | hv208 | Household possesses TV | X |
| | HR | hv210 | Household possesses bicycle | X |
| | HR | hv206 | Household possesses electricity | X |
| Type of Sanitation Facility | HR | hv205 | Household has improved sanitation facility | X |
| | HR | hv225 | Household does not share toilet with other households | X |
| Wealth Quintiles | HR/WI | hv271 | Household's wealth index factor (percentile)<br>*Note only earlier DHS datasets had the WI database, other DHS datasets had this variable in the HR database* | X |
| Other | BR | v025 | Household in urban area | X |
| **3) Respondent's characteristics** | | | | |
| Educational Attainment of Household Members | BR | v133 | Mother's years of education | |
| Employment and Occupation | BR | v731 | Mother works or worked in past year | X |
| Literacy | BR | v155, v106 | Mother is literate | |
| Other | BR | v012 | Mother's age | X |
| **4) Marriage and Sexual Activity** | | | | |
| Age at First Marriage | BR | v511 | Mother got married younger than 18 years old | |
| Age at First Sexual Intercourse | BR | v531 | Mother had first intercourse before 15 years old | |
| Current Marital Status | BR | v501 | Mother has partner | X |
| **5) Fertility** | | | | |
| Age at First Birth | BR | v212 | Mother had first child before 18 years old | |
| Children Ever Born and Living | BR | v201 | Mother's number of children ever born | X |
| **6) Fertility Preferences** | | | | |
| Fertility Planning | BR | m10 | Mother wanted pregnancy (wanted before or during pregnancy) | |
| **7) Family Planning** | | | | |
| Contact of Nonusers with Family Planning Providers | BR | v394 | Mother visited health facility in last 12 months | |
| Current Use of Contraceptive Methods | IR | v313 | Mother uses modern contraceptive method | |
| | IR | v313, v361, v362 | Mother has used or plans to use contraceptive methods | |
| Decision Making about Family Planning | BR | V632, v632a | Mother participates in family planning decisions | |
| Exposure to Family Planning Messages | BR | v384a | Mother heard about family planning in the radio (last months) | |
| | BR | v384b | Mother heard about family planning in TV (last months) | |
| | BR | v384c | Mother read about family planning in newspaper (last months) | |
| Informed Choice | BR | V3a02, v3a03, v3a04 | Mother was told side-effects when getting contraceptives | |

| | | | |
|---|---|---|---|
| Knowledge of Contraceptive Methods | IR | V304_01, 02, 03, 05, 06, 07, 08, 09, 11 | Mother’s number of known contraceptive methods (out of 9) |
| Knowledge of the Fertile Period | BR | v217 | Mothers knows her ovulatory cycle |
| **9) Reproductive Health** | | | |
| Antenatal Care | BR | m14 | Mother had at least 1 antenatal visit during pregnancy |
| | BR | m42c | Mother had blood pressure taken during pregnancy |
| | BR | m42d | Mother had urine sample taken during pregnancy |
| | BR | m42e | Mother had blood sample taken during pregnancy |
| | BR | m42d, e | Mother had blood or urine sample taken during pregnancy |
| Assistance during Delivery | BR | m3a, b, c | Mother assisted during delivery by skilled provider |
| Place of Delivery | BR | m15 | Mother delivered at home |
| Problems in Accessing Health Care | BR | v467c | Mother says money is a problem to access healthcare when sick |
| | BR | v467d | Mother says distance is a problem to access healthcare when sick |
| | BR | v467b | Mother says getting permission to go is a problem to access healthcare when sick |
| **10) Child health** | | | |
| Knowledge of ORS packets | BR | v416 | Mother knows Oral Rehydration Salts packets |
| **11) Nutrition of Children & Adults** | | | |
| Initial Breastfeeding | BR | m4 | Mother ever breastfed her child |
| | BR | m34/m4 | Mother breastfed her child the day it was born |
| **15) Women’s Empowerment** | | | |
| Attitude towards Wife Beating | BR | v744a | Mother justifies being beaten for going out without permission |
| | BR | v744b | Mother justifies being beaten for neglecting a child |
| | BR | v744c | Mother justifies being beaten for arguing with partner |
| | BR | v744d | Mother justifies being beaten for not having sex |
| | BR | v744e | Mother justifies being beaten for burning food |
| | BR | v744a-e | Mother justifies being beaten |
| Participation in Decision Making | BR | v743a, v502 | Mother decides on her own health |
| | BR | v743b, v502 | Mother decides on large household purchases |
| | BR | v743d, v502 | Mother decides on visits to family or relatives |

*Table 2: Computing variables from DHS data*

| Variables | Calculation |
|---|---|
| **2) Population & Housing** | |
| Household's number of residents | log(hv012).<br>if hv012==0, then we set hv012 as 1 (so we can compute the log) |
| Household has improved source of drinking water | TRUE if hv201 is any of the following: 11, 12, 13, 31, 32, 33, 51, 61, 71, "piped into dwelling", "piped into house/yard/plot", "piped to yard/plot", "piped into yard/plot", "public tap/standpipe", "public/nieghbor's tap", "communal tap", "piped to neighbor", "piped to neighbour", "tube well or borehole", "tubewell in yard/plot", "public/neighbor's tubewell", "protected well", "protected well in yard/plot", "protected public well", "protected spring", "stone tap/dhara", "rainwater", "tanker truck", "cart with small tank", "bottled water" |
| Household possesses radio | TRUE if hv207 is "yes" or 1 |
| Household possesses TV | TRUE if hv208 is "yes" or 1 |
| Household possesses bicycle | TRUE if hv210 is "yes" or 1 |
| Household possesses electricity | TRUE if hv206 is "yes" or 1 |
| Household has improved sanitation facility | TRUE if hv201 is any of the following: 11, 12, 22, "flush to piped sewer system", "flush toilet", "flush to septic tank", "flush to pit latrine", "flush, don't know where", "flush to somewhere else", "ventilated improved pit latrine (vip)", "ventilated/improved pit latrine", "vip", "pit latrine with slab", "traditional pit toilet", "composting toilet".<br>*DHS recommendations say that "flush to somewhere else" is not considered an improved sanitation facility, but we had to consider it as improved since earlier DHS datasets did not make this distinction and only have "flush toilet"; the number of samples with "flush to somewhere else" is minimal so it is not a big issue.* |
| Household does not share toilet with other households | TRUE if hv225 is "no" or 0 |
| Household's wealth index factor (percentile) | Percentile of hv271 when compared to all samples (for same year-country DHS) |
| Household in urban area | TRUE if v025 is"urban" |
| **3) Respondent's characteristics** | |
| Mother's years of education | log(1+v133).<br>if v133>90 or v133==NA, then we set v133 as 0 |
| Mother works or worked in past year | TRUE if v731 is any of the following: 1, 2, 3, "in the past year", "currently working", "have a job, but on leave last 7 days" |
| Mother is literate | TRUE if v106 is 3 or "higher", or if v155 is any of the following: 1, 2, "able to read only parts of sentence", "able to read whole sentence" |
| Mother's age | log(v012) |
| **4) Marriage and Sexual Activity** | |
| Mother got married younger than 18 years old | TRUE if v511>0 and v511<18.<br>NA values of v511 are considered to be 0 (never married) |
| Mother had first intercourse before 15 years old | TRUE if v531>0 and v531<15.<br>NA values of v511 are considered to be 0 |
| Mother has partner | TRUE if v501 is any of the following: "married", "living together", "living with partner" |
| **5) Fertility** | |
| Mother had first child before 18 years old | TRUE if v212<18 |
| Mother's number of children ever born | log(v212) |
| **6) Fertility Preferences** | |
| Mother wanted pregnancy (wanted before or during pregnancy) | TRUE if m10 is any of the following: 1, 2, "then", "later" |
| **7) Family Planning** | |
| Mother visited health facility in last 12 months | TRUE if v394 is 1 or "yes" |

| | |
|---|---|
| Mother uses modern contraceptive method | TRUE if v313 is "modern method" |
| Mother has used or plans to use contraceptive methods | TRUE if v313 is "modern method", or either v361 or v362 is any of the following: 1, 2, 3, "in next 12 months", "use later", "unsure about timing ", "currently using", "used since last birth", "used before last birth" |
| Mother participates in family planning decisions | TRUE if either v632 or v632a is any of the following: 1, 3, "mainly respondent", "joint decision".<br>*Note some DHS datasets did not have variable v632a and was not considered in such case* |
| Mother heard about family planning in the radio | TRUE if v384a is 1 or "yes" |
| Mother heard about family planning in TV | TRUE if v384b is 1 or "yes" |
| Mother read about family planning in newspaper | TRUE if v384c is 1 or "yes" |
| Mother was told side-effects when getting contraceptives | TRUE if either v3a02, v3a03, or v3a04 is either 1 or "yes"<br>*Note some DHS datasets did not have variable v3a02 and was not considered in such case* |
| Mother’s number of known contraceptive methods | Count the number of variables among v304_01, 02, 03, 05, 06, 07, 08, 09, 11, that have any of the following values: 1, 7, "yes", "not in contraception table, but as a current method".<br>*Note we only considered contraceptive methods included in all DHS datasets* |
| Mothers knows her ovulatory cycle | TRUE if v217 is any of the following: 1, 2, 3, 4, 5, "during her period", "after period ended", "middle of the cycle", "before period begins", "at any time" |
| **9) Reproductive Health** | |
| Mother had at least 1 antenatal visit during pregnancy | TRUE if m14>0.<br>NA values or values larger than 90 of m14 are considered to be 0 |
| Mother had blood pressure taken during pregnancy | TRUE if m42c is 1 or "yes" |
| Mother had urine sample taken during pregnancy | TRUE if m42d is 1 or "yes" |
| Mother had blood sample taken during pregnancy | TRUE if m42e is 1 or "yes" |
| Mother had blood or urine sample taken during pregnancy | TRUE if either m42d or m42e is 1 or "yes" |
| Mother assisted during delivery by skilled provider | TRUE if m3a or m3b are any of the following: 1, "yes", "yes: doctor", "yes: nurse/midwife".<br>*Note m3a and b correspond to “doctor” and “nurse/midwife” respectively for all but one DHS datasets. For Senegal 2019 DHS, m3b only corresponds to “nurse”, so we also consider m3c that corresponds to “midwife” in that case.* |
| Mother delivered at home | TRUE if m15 is any of the following: 11, 12, "respondent's home", "respondents home", "other home" |
| Mother says money is a problem to access healthcare when sick | TRUE if v467c is any of the following: 1, "big problem", "a big problem" |
| Mother says distance is a problem to access healthcare when sick | TRUE if v467d is any of the following: 1, "big problem", "a big problem" |
| Mother says getting permission to go is a problem to access healthcare when sick | TRUE if v467b is any of the following: 1, "big problem", "a big problem" |
| **10) Child health** | |
| Mother knows Oral Rehydration Salts packets | TRUE if v416 is any of the following: 1, 2, "used ors", "heard of ors" |
| **11) Nutrition of Children & Adults** | |
| Mother ever breastfed her child | TRUE if m4 is any of the following: 0 to 59, 95, "ever breastfed, not currently breastfeeding", "still breastfeeding" |
| Mother breastfed her child the | TRUE if m34<124 and m4 is any of the following: 0 to 59, 95, "ever breastfed, not |

| | |
|---|---|
| day it was born | currently breastfeeding", "still breastfeeding" |
| **15) Women's Empowerment** | |
| Mother justifies being beaten for going out without permission | TRUE if v744a is 1 or "yes" |
| Mother justifies being beaten for neglecting a child | TRUE if v744b is 1 or "yes" |
| Mother justifies being beaten for arguing with partner | TRUE if v744c is 1 or "yes" |
| Mother justifies being beaten for not having sex | TRUE if v744d is 1 or "yes" |
| Mother justifies being beaten for burning food | TRUE if v744e is 1 or "yes" |
| Mother justifies being beaten | TRUE if any of v744a, b, c, d, or e is 1 or "yes" |
| Mother decides on her own health | TRUE if v743a is any of the following: 1, 2, 3, "respondent alone", "respondent and husband/partner", "respondent and other person". Also TRUE if v502 is NOT one of the following: "currently married", "currently in union/living with a man". *Note later DHS datasets only ask this question to married woman, so that is why we added the v502 criteria.* |
| Mother decides on large household purchases | TRUE if v743b is any of the following: 1, 2, 3, "respondent alone", "respondent and husband/partner", "respondent and other person". Also TRUE if v502 is NOT one of the following: "currently married", "currently in union/living with a man". *Note later DHS datasets only ask this question to married woman, so that is why we added the v502 criteria.* |
| Mother decides on visits to family or relatives | TRUE if v743d is any of the following: 1, 2, 3, "respondent alone", "respondent and husband/partner", "respondent and other person". Also TRUE if v502 is NOT one of the following: "currently married", "currently in union/living with a man". *Note later DHS datasets only ask this question to married woman, so that is why we added the v502 criteria.* |

*Data sources: Various DHS [1].*
*Note some DHS datasets and variables had numbers instead of labels for their categorical values.*

### B: Additional results

*Table 3: VIF of control variables*

| Variables | Nepal | | Senegal | | Zambia | |
|---|---|---|---|---|---|---|
| | 2001 | 2016 | 2005 | 2019 | 2001-02 | 2018 |
| **2) Population & Housing** | | | | | | |
| Household's number of residents | 1.25 | 1.28 | 1.16 | 1.24 | 1.42 | 1.40 |
| Household has improved source of drinking water | 1.17 | 1.08 | 1.40 | 1.28 | 1.65 | 1.25 |
| Household possesses radio | 1.34 | 1.04 | 1.08 | 1.08 | 1.64 | 1.45 |
| Household possesses TV | 1.55 | 1.56 | 2.55 | 2.97 | 2.27 | 3.04 |
| Household possesses bicycle | 1.64 | 1.54 | 1.19 | 1.15 | 1.17 | 1.25 |
| Household possesses electricity | 1.82 | 1.20 | 3.22 | 2.86 | 2.57 | 3.47 |
| Household has improved sanitation facility | 3.35 | 2.11 | 1.64 | 1.87 | 1.93 | 1.40 |
| Household does not share toilet with other households | 2.67 | 1.88 | 1.31 | 1.29 | 1.19 | 1.14 |
| Household's wealth index factor (percentile) | 2.87 | 2.28 | 4.87 | 6.92 | 3.51 | 3.88 |
| Household in urban area | 1.35 | 1.07 | 1.84 | 1.90 | 2.54 | 2.74 |
| **3) Respondent's characteristics** | | | | | | |
| Mother works or worked in past year | 1.22 | 1.24 | 1.09 | 1.07 | 1.11 | 1.05 |
| Mother's age | 2.94 | 2.03 | 2.65 | 2.59 | 3.98 | 3.35 |
| **4) Marriage and Sexual Activity** | | | | | | |
| Mother has partner | 1.03 | 1.03 | 1.06 | 1.06 | 1.22 | 1.18 |
| **5) Fertility** | | | | | | |
| Mother's number of children ever born | 3.11 | 2.22 | 2.70 | 2.76 | 4.41 | 3.78 |

*Data sources: Various DHS [1].*
*Each value corresponds to the VIF of the variable when compared to all other control variables*

*Table 4: Statistical significance of OLR coefficients of control variables*

| | Nepal | | Senegal | | Zambia | | Count of coefficients not containing zero | |
|---|---|---|---|---|---|---|---|---|
| **Variables** | **2001** | **2016** | **2005** | **2019** | **2001-02** | **2018** | **(90% CI)** | **[95% CI]** |
| **2) Population & Housing** | | | | | | | | |
| Household's number of residents | -0.0986<br>(-0.20 to 0.00)<br>[-0.22 to 0.02] | -0.1759<br>(-0.33 to -0.02)*<br>[-0.36 to 0.01] | 0.0984<br>(0.00 to 0.20)*<br>[-0.02 to 0.22] | 0.2010<br>(0.05 to 0.35)*<br>[0.02 to 0.38]* | -0.0300<br>(-0.18 to 0.12)<br>[-0.21 to 0.15] | 0.0327<br>(-0.11 to 0.18)<br>[-0.14 to 0.20] | 3 | 1 |
| Household has improved source of drinking water | 0.2367<br>(0.02 to 0.45)*<br>[-0.02 to 0.49] | -0.1858<br>(-0.83 to 0.46)<br>[-0.96 to 0.59] | -0.1759<br>(-0.33 to -0.02)*<br>[-0.36 to 0.01] | 0.3822<br>(0.08 to 0.69)*<br>[0.02 to 0.75]* | 0.7480<br>(0.55 to 0.95)*<br>[0.51 to 0.99]* | 0.1597<br>(-0.02 to 0.34)<br>[-0.06 to 0.38] | 4 | 2 |
| Household possesses radio | 0.8611<br>(0.66 to 1.06)*<br>[0.62 to 1.10]* | 0.7795<br>(0.44 to 1.12)*<br>[0.38 to 1.18]* | 0.2604<br>(0.03 to 0.49)*<br>[-0.01 to 0.53] | 0.3091<br>(0.07 to 0.55)*<br>[0.02 to 0.60]* | 0.2548<br>(0.06 to 0.45)*<br>[0.02 to 0.49]* | 0.1741<br>(0.00 to 0.35)*<br>[-0.03 to 0.38] | 6 | 4 |
| Household possesses TV | 0.9284<br>(0.56 to 1.29)*<br>[0.50 to 1.36]* | 0.6188<br>(0.36 to 0.88)*<br>[0.31 to 0.93]* | 0.3561<br>(0.21 to 0.50)*<br>[0.18 to 0.53]* | 0.5437<br>(0.32 to 0.77)*<br>[0.27 to 0.81]* | 0.7918<br>(0.49 to 1.09)*<br>[0.43 to 1.15]* | 0.2773<br>(0.09 to 0.46)*<br>[0.05 to 0.50]* | 6 | 6 |
| Household possesses bicycle | 0.2185<br>(0.01 to 0.43)*<br>[-0.03 to 0.47] | -0.5041<br>(-0.75 to -0.26)*<br>[-0.80 to -0.21]* | 0.0763<br>(-0.12 to 0.27)<br>[-0.15 to 0.31] | -0.2405<br>(-0.55 to 0.07)<br>[-0.61 to 0.13] | 0.0352<br>(-0.17 to 0.24)<br>[-0.21 to 0.28] | 0.0708<br>(-0.10 to 0.25)<br>[-0.14 to 0.28] | 2 | 1 |
| Household possesses electricity | 0.8123<br>(0.53 to 1.10)*<br>[0.48 to 1.15]* | 0.3751<br>(0.01 to 0.74)*<br>[-0.06 to 0.81] | 0.3140<br>(0.17 to 0.46)*<br>[0.14 to 0.49]* | 0.4206<br>(0.19 to 0.66)*<br>[0.14 to 0.70]* | 0.5897<br>(0.28 to 0.90)*<br>[0.22 to 0.96]* | 0.1736<br>(-0.02 to 0.37)<br>[-0.06 to 0.40] | 5 | 4 |
| Household has improved sanitation facility | 1.4301<br>(1.14 to 1.72)*<br>[1.08 to 1.78]* | 0.8587<br>(0.59 to 1.13)*<br>[0.54 to 1.18]* | 0.1952<br>(0.05 to 0.34)*<br>[0.02 to 0.37]* | 0.5765<br>(0.34 to 0.82)*<br>[0.29 to 0.86]* | 0.5564<br>(0.25 to 0.86)*<br>[0.19 to 0.92]* | 0.0596<br>(-0.11 to 0.23)<br>[-0.15 to 0.26] | 5 | 5 |
| Household does not share toilet with other households | 1.3717<br>(1.02 to 1.72)*<br>[0.95 to 1.79]* | 0.5930<br>(0.35 to 0.84)*<br>[0.30 to 0.89]* | 0.2590<br>(0.12 to 0.40)*<br>[0.09 to 0.43]* | 0.6109<br>(0.37 to 0.85)*<br>[0.33 to 0.90]* | -0.0276<br>(-0.23 to 0.17)<br>[-0.27 to 0.21] | 0.2213<br>(0.05 to 0.39)*<br>[0.02 to 0.43]* | 5 | 5 |
| Household's wealth index factor (percentile) | 0.7893<br>(0.61 to 0.96)*<br>[0.58 to 1.00]* | 0.2524<br>(0.04 to 0.47)*<br>[-0.01 to 0.51] | 0.1006<br>(-0.02 to 0.22)<br>[-0.04 to 0.24] | 0.5628<br>(0.37 to 0.75)*<br>[0.34 to 0.79]* | 0.5351<br>(0.36 to 0.71)*<br>[0.33 to 0.74]* | 0.2809<br>(0.13 to 0.43)*<br>[0.10 to 0.46]* | 5 | 4 |
| Household in urban area | 0.4960<br>(0.08 to 0.91)*<br>[0.00 to 0.99] | 0.1043<br>(-0.14 to 0.35)<br>[-0.19 to 0.40] | 0.0539<br>(-0.09 to 0.20)<br>[-0.12 to 0.23] | 0.2916<br>(0.05 to 0.53)*<br>[0.01 to 0.57]* | 0.4789<br>(0.25 to 0.71)*<br>[0.21 to 0.75]* | 0.1785<br>(0.00 to 0.36)<br>[-0.04 to 0.40] | 3 | 2 |
| **3) Respondent's characteristics** | | | | | | | | |
| Mother works or worked in past year | -0.0743<br>(-0.33 to 0.18)<br>[-0.37 to 0.22] | 0.4708<br>(0.22 to 0.72)*<br>[0.18 to 0.77]* | -0.0271<br>(-0.17 to 0.12)<br>[-0.20 to 0.14] | 0.1259<br>(-0.10 to 0.35)<br>[-0.14 to 0.39] | -0.2325<br>(-0.44 to -0.03)*<br>[-0.47 to 0.01] | -0.1591<br>(-0.33 to 0.01)<br>[-0.37 to 0.05] | 2 | 1 |
| Mother's age | -0.4112<br>(-0.56 to -0.27)*<br>[-0.58 to -0.24]* | -0.0746<br>(-0.25 to 0.10)<br>[-0.28 to 0.14] | 0.0167<br>(-0.09 to 0.12)<br>[-0.11 to 0.14] | -0.1119<br>(-0.30 to 0.08)<br>[-0.34 to 0.12] | -0.2051<br>(-0.34 to -0.07)*<br>[-0.37 to -0.04]* | -0.0643<br>(-0.20 to 0.07)<br>[-0.23 to 0.10] | 2 | 2 |
| **4) Marriage and Sexual Activity** | | | | | | | | |
| Mother has partner | 0.6179<br>(-0.28 to 1.52)<br>[-0.45 to 1.69] | 2.1798<br>(0.54 to 3.82)*<br>[0.23 to 4.13]* | 0.2132<br>(-0.11 to 0.53)<br>[-0.17 to 0.60] | -0.3793<br>(-0.95 to 0.19)<br>[-1.06 to 0.30] | 0.0583<br>(-0.20 to 0.31)<br>[-0.24 to 0.36] | 0.0642<br>(-0.15 to 0.27)<br>[-0.19 to 0.32] | 1 | 1 |
| **5) Fertility** | | | | | | | | |
| Mother's number of children ever born | -0.3366<br>(-0.44 to -0.24)*<br>[-0.46 to -0.22]* | -0.7075<br>(-0.93 to -0.48)*<br>[-0.98 to -0.44]* | -0.1438<br>(-0.23 to -0.05)*<br>[-0.25 to -0.04]* | -0.3690<br>(-0.52 to -0.22)*<br>[-0.55 to -0.19]* | -0.2594<br>(-0.38 to -0.13)*<br>[-0.41 to -0.11]* | -0.1393<br>(-0.25 to -0.03)*<br>[-0.27 to -0.01]* | 6 | 6 |

*Data sources: Various DHS [1].*
*Coefficients were obtained from ordinal logistic regressions; children groups LV, MV, and 8V were used as ordinal categories and each control variable was tested separately.*
** denotes CIs of coefficients not containing zero. Values represent the average value of the coefficient, parentheses "()" show 90% CI, brackets "[]" show 95% CI.*

*Table 5: Average values of control variables, by DHS dataset and vaccine group*

| Variables | Vaccine group | Nepal | | Senegal | | Zambia | | Count of 8V-LV differences not containing zero (95% CI) |
|---|---|---|---|---|---|---|---|---|
| | | 2001† | 2016‡ | 2005† | 2019‡ | 2001-02† | 2018‡ | |
| **2) Population & Housing** | | | | | | | | |
| Household's number of residents | LV | 6.3 (5.9 to 6.8) | 6.9 (5.7 to 8.3) | 11.6 (10.6 to 12.7) | 11.3 (9.9 to 13.1) | 7.1 (6.3 to 8.1) | 6.4 (5.9 to 7.1) | |
| | MV | 7.0 (6.7 to 7.4) | 6.1 (5.7 to 6.6) | 13.2 (12.7 to 13.8) | 13.0 (11.9 to 14.2) | 6.3 (6.0 to 6.7) | 6.0 (5.8 to 6.3) | |
| | 8V | 6.6 (6.3 to 6.8) | 5.8 (5.6 to 6.0) | 13.1 (12.7 to 13.5) | 13.4 (12.9 to 14.0) | 6.3 (6.1 to 6.5) | 6.2 (6.0 to 6.3) | |
| | 8V-LV | 0.3 (-0.3 to 0.8) | -1.1 (-2.5 to 0.1) | 1.5 (0.3 to 2.6)* | 2.1 (0.3 to 3.7)* | -0.8 (-1.8 to 0.0) | -0.3 (-0.9 to 0.3) | 2 |
| Household has improved source of drinking water | LV | 53% (42% to 63%) | 100% (100% to 100%) | 66% (58% to 74%) | 71% (61% to 82%) | 28% (17% to 39%) | 54% (44% to 66%) | |
| | MV | 76% (71% to 80%) | 96% (92% to 98%) | 70% (66% to 73%) | 86% (81% to 91%) | 39% (34% to 44%) | 69% (64% to 73%) | |
| | 8V | 74% (71% to 77%) | 96% (95% to 97%) | 65% (62% to 68%) | 87% (84% to 89%) | 55% (52% to 58%) | 69% (67% to 71%) | |
| | 8V-LV | 21% (11% to 33%)* | -4% (-5% to -3%)* | -1% (-9% to 7%) | 15% (5% to 26%)* | 27% (16% to 38%)* | 15% (4% to 25%)* | 5 |
| Household possesses radio | LV | 30% (22% to 40%) | 17% (7% to 29%) | 83% (76% to 89%) | 60% (48% to 71%) | 36% (25% to 48%) | 40% (30% to 51%) | |
| | MV | 29% (24% to 34%) | 13% (9% to 19%) | 89% (87% to 91%) | 69% (62% to 75%) | 40% (35% to 45%) | 45% (40% to 50%) | |
| | 8V | 50% (46% to 53%) | 27% (24% to 30%) | 90% (88% to 92%) | 72% (69% to 75%) | 45% (42% to 48%) | 48% (46% to 51%) | |
| | 8V-LV | 20% (9% to 28%)* | 9% (-3% to 21%) | 7% (1% to 14%)* | 13% (1% to 25%)* | 9% (-3% to 21%) | 9% (-2% to 19%) | 3 |
| Household possesses TV | LV | 2% (0% to 6%) | 33% (19% to 48%) | 28% (21% to 35%) | 38% (27% to 48%) | 13% (6% to 20%) | 23% (15% to 33%) | |
| | MV | 7% (5% to 10%) | 34% (27% to 41%) | 41% (37% to 45%) | 56% (50% to 63%) | 10% (7% to 13%) | 30% (26% to 35%) | |
| | 8V | 14% (11% to 16%) | 49% (45% to 52%) | 47% (44% to 50%) | 64% (61% to 67%) | 21% (18% to 23%) | 35% (32% to 37%) | |
| | 8V-LV | 12% (7% to 15%)* | 15% (1% to 30%)* | 19% (10% to 27%)* | 27% (15% to 37%)* | 8% (-1% to 15%) | 11% (1% to 20%)* | 5 |
| Household possesses bicycle | LV | 10% (4% to 17%) | 64% (50% to 76%) | 14% (8% to 19%) | 19% (12% to 30%) | 35% (25% to 48%) | 41% (32% to 52%) | |
| | MV | 31% (26% to 36%) | 48% (40% to 55%) | 16% (14% to 18%) | 15% (10% to 20%) | 35% (30% to 40%) | 39% (34% to 44%) | |
| | 8V | 30% (27% to 33%) | 39% (36% to 43%) | 16% (14% to 18%) | 13% (11% to 16%) | 36% (33% to 39%) | 41% (39% to 44%) | |
| | 8V-LV | 20% (13% to 26%)* | -25% (-39% to -10%)* | 3% (-4% to 9%) | -7% (-16% to 2%) | 0% (-12% to 11%) | 0% (-11% to 10%) | 2 |
| Household possesses electricity | LV | 9% (3% to 16%) | 81% (69% to 93%) | 22% (17% to 29%) | 47% (36% to 58%) | 19% (10% to 28%) | 20% (11% to 29%) | 4 |
| | MV | 11% (8% to 15%) | 87% (83% to 92%) | 42% (38% to 46%) | 67% (60% to 74%) | 8% (5% to 11%) | 29% (24% to 33%) | |
| | 8V | 22% (19% to 25%) | 90% (87% to 92%) | 45% (43% to 48%) | 70% (67% to 73%) | 17% (15% to 20%) | 30% (28% to 32%) | |

| | | | | | | | | |
|---|---|---|---|---|---|---|---|---|
| | 8V-LV | 13%<br>(6% to 19%)* | 8%<br>(-3% to 21%) | 23%<br>(15% to 30%)* | 23%<br>(12% to 34%)* | -2%<br>(-11% to 7%) | 10%<br>(1% to 19%)* | |
| Household has improved sanitation facility | LV | 4%<br>(0% to 9%) | 67%<br>(52% to 79%) | 31%<br>(24% to 39%) | 52%<br>(42% to 64%) | 20%<br>(12% to 30%) | 37%<br>(26% to 46%) | 5 |
| | MV | 11%<br>(8% to 14%) | 63%<br>(56% to 70%) | 42%<br>(38% to 46%) | 67%<br>(61% to 74%) | 8%<br>(6% to 11%) | 53%<br>(47% to 57%) | |
| | 8V | 30%<br>(27% to 33%) | 81%<br>(78% to 83%) | 44%<br>(41% to 47%) | 75%<br>(72% to 78%) | 17%<br>(15% to 20%) | 50%<br>(48% to 53%) | |
| | 8V-LV | 26%<br>(20% to 30%)* | 14%<br>(2% to 29%)* | 14%<br>(5% to 21%)* | 23%<br>(11% to 34%)* | -3%<br>(-12% to 6%) | 14%<br>(4% to 24%)* | |
| Household does not share toilet with other households | LV | 0%<br>(0% to 2%) | 40%<br>(26% to 55%) | 42%<br>(34% to 50%) | 48%<br>(38% to 58%) | 45%<br>(33% to 57%) | 49%<br>(40% to 60%) | 4 |
| | MV | 7%<br>(5% to 10%) | 48%<br>(41% to 55%) | 56%<br>(52% to 59%) | 67%<br>(61% to 74%) | 37%<br>(32% to 43%) | 48%<br>(43% to 53%) | |
| | 8V | 20%<br>(17% to 23%) | 61%<br>(57% to 64%) | 59%<br>(56% to 62%) | 75%<br>(72% to 77%) | 38%<br>(35% to 42%) | 54%<br>(51% to 57%) | |
| | 8V-LV | 19%<br>(16% to 22%)* | 19%<br>(5% to 34%)* | 17%<br>(8% to 25%)* | 27%<br>(16% to 37%)* | -7%<br>(-19% to 5%) | 4%<br>(-6% to 15%) | |
| Household's wealth index factor (percentile) | LV | 33%<br>(28% to 38%) | 51%<br>(43% to 59%) | 44%<br>(40% to 49%) | 35%<br>(29% to 42%) | 49%<br>(41% to 55%) | 45%<br>(38% to 51%) | 5 |
| | MV | 44%<br>(41% to 47%) | 50%<br>(46% to 54%) | 54%<br>(52% to 57%) | 55%<br>(51% to 60%) | 48%<br>(45% to 51%) | 51%<br>(48% to 54%) | |
| | 8V | 54%<br>(52% to 55%) | 54%<br>(52% to 56%) | 54%<br>(52% to 56%) | 59%<br>(57% to 61%) | 57%<br>(55% to 59%) | 55%<br>(53% to 56%) | |
| | 8V-LV | 21%<br>(15% to 26%)* | 3%<br>(-5% to 12%) | 9%<br>(4% to 14%)* | 24%<br>(17% to 30%)* | 8%<br>(1% to 16%)* | 10%<br>(3% to 16%)* | |
| Household in urban area | LV | 2%<br>(0% to 7%) | 45%<br>(31% to 60%) | 28%<br>(20% to 35%) | 25%<br>(17% to 35%) | 22%<br>(13% to 32%) | 28%<br>(18% to 37%) | 3 |
| | MV | 5%<br>(3% to 8%) | 55%<br>(47% to 62%) | 40%<br>(36% to 43%) | 36%<br>(29% to 43%) | 23%<br>(18% to 27%) | 34%<br>(29% to 38%) | |
| | 8V | 8%<br>(6% to 9%) | 55%<br>(52% to 58%) | 38%<br>(36% to 41%) | 39%<br>(36% to 43%) | 32%<br>(29% to 35%) | 36%<br>(34% to 38%) | |
| | 8V-LV | 5%<br>(0% to 8%)* | 10%<br>(-6% to 24%) | 11%<br>(3% to 18%)* | 14%<br>(4% to 24%)* | 10%<br>(-1% to 19%) | 9%<br>(-1% to 18%) | |
| **3) Respondent's characteristics** | | | | | | | | |
| Mother works or worked in past year | LV | 96%<br>(91% to 99%) | 45%<br>(31% to 60%) | 36%<br>(29% to 44%) | 48%<br>(38% to 58%) | 68%<br>(58% to 78%) | 48%<br>(38% to 59%) | 1 |
| | MV | 80%<br>(76% to 84%) | 49%<br>(42% to 56%) | 46%<br>(42% to 49%) | 49%<br>(41% to 55%) | 64%<br>(59% to 69%) | 61%<br>(56% to 66%) | |
| | 8V | 83%<br>(81% to 86%) | 60%<br>(56% to 63%) | 43%<br>(40% to 46%) | 52%<br>(48% to 55%) | 60%<br>(56% to 63%) | 54%<br>(51% to 56%) | |
| | 8V-LV | -13%<br>(-17% to -7%)* | 15%<br>(-1% to 30%) | 7%<br>(-1% to 14%) | 3%<br>(-8% to 15%) | -9%<br>(-19% to 3%) | 6%<br>(-5% to 17%) | |
| Mother's age | LV | 29.5<br>(28.1 to 31.0) | 27.4<br>(25.7 to 29.0) | 27.9<br>(26.7 to 29.1) | 29.9<br>(28.4 to 31.3) | 30.1<br>(28.3 to 31.9) | 28.3<br>(26.8 to 30.1) | 3 |
| | MV | 27.3<br>(26.5 to 27.9) | 25.0<br>(24.2 to 25.8) | 28.2<br>(27.6 to 28.7) | 29.6<br>(28.6 to 30.5) | 27.2<br>(26.4 to 28.0) | 28.1<br>(27.4 to 28.8) | |
| | 8V | 26.1<br>(25.7 to 26.5) | 25.3<br>(24.9 to 25.6) | 28.1<br>(27.7 to 28.5) | 29.2<br>(28.8 to 29.6) | 26.7<br>(26.3 to 27.1) | 27.8<br>(27.4 to 28.2) | |
| | 8V-LV | -3.5<br>(-4.9 to -2.0)* | -2.1<br>(-3.7 to -0.4)* | 0.2<br>(-1.0 to 1.5) | -0.7<br>(-2.2 to 0.8) | -3.3<br>(-5.2 to -1.5)* | -0.5<br>(-2.2 to 1.1) | |
| **4) Marriage and Sexual Activity** | | | | | | | | |

| | | | | | | | | |
|---|---|---|---|---|---|---|---|---|
| Mother has partner | LV | 100% (100% to 100%) | 100% (100% to 100%) | 91% (87% to 95%) | 97% (94% to 100%) | 78% (68% to 88%) | 78% (68% to 86%) | |
| | MV | 98% (97% to 99%) | 99% (97% to 100%) | 95% (93% to 97%) | 96% (93% to 99%) | 83% (79% to 86%) | 79% (75% to 82%) | |
| | 8V | 99% (99% to 100%) | 100% (100% to 100%) | 95% (94% to 96%) | 95% (93% to 96%) | 83% (80% to 85%) | 80% (77% to 82%) | |
| | 8V-LV | -1% (-1% to 0%)* | 0% (0% to 0%)* | 4% (-1% to 9%) | -3% (-6% to 1%) | 3% (-6% to 15%) | 2% (-7% to 12%) | 2 |
| **5) Fertility** | | | | | | | | |
| Mother's number of children ever born | LV | 4.3 (3.8 to 4.8) | 3.3 (2.7 to 3.9) | 4.2 (3.8 to 4.6) | 4.0 (3.6 to 4.5) | 5.2 (4.5 to 5.9) | 4.0 (3.5 to 4.7) | |
| | MV | 3.5 (3.3 to 3.7) | 2.6 (2.3 to 2.8) | 3.9 (3.7 to 4.1) | 3.8 (3.4 to 4.1) | 3.9 (3.6 to 4.1) | 3.7 (3.4 to 3.9) | 6 |
| | 8V | 3.0 (2.9 to 3.1) | 2.2 (2.1 to 2.3) | 3.6 (3.5 to 3.8) | 3.3 (3.2 to 3.4) | 3.6 (3.4 to 3.7) | 3.4 (3.3 to 3.5) | |
| | 8V-LV | -1.3 (-1.8 to -0.8)* | -1.1 (-1.7 to -0.5)* | -0.6 (-1.0 to -0.2)* | -0.7 (-1.2 to -0.3)* | -1.6 (-2.3 to -0.9)* | -0.6 (-1.3 to 0.0)* | |

*Data sources: Various DHS [1].*

*† denotes DHS surveys where LV consists of one-year-old children with 0-2 vaccines and MV of 3-7 vaccines.*

*‡ denotes DHS surveys where LV consists of one-year-old children with 0-4 vaccines and MV of 5-7 vaccines.*

** denotes CIs not containing zero. Values correspond to the median of the averages of groups LV, MV, 8V, and 8V-LV differences. Value ranges in parenthesis correspond to the 95% CI obtained by a weighted bootstrap method of 1000 replications.*

*Table 6: Statistical significance of OLR coefficients of tested variables*

| Variables | Nepal | | Senegal | | Zambia | | Count of coefficients not containing zero | |
|---|---|---|---|---|---|---|---|---|
| | 2001 | 2016 | 2005 | 2019 | 2001-02 | 2018 | (90% CI) | [95% CI] |
| **3) Respondent's characteristics** | | | | | | | | |
| Mother’s years of education | 0.7543<br>(0.55 to 0.96)*<br>[0.51 to 0.99]* | 0.3755<br>(0.01 to 0.74)*<br>[-0.06 to 0.81] | 0.5258<br>(0.41 to 0.64)*<br>[0.39 to 0.66]* | 0.2157<br>(-0.03 to 0.46)<br>[-0.08 to 0.51] | 0.2604<br>(0.13 to 0.39)*<br>[0.10 to 0.42]* | 0.2057<br>(0.12 to 0.29)*<br>[0.10 to 0.31]* | 5 | 4 |
| Mother is literate | 1.1404<br>(0.87 to 1.41)*<br>[0.81 to 1.47]* | 0.1082<br>(-0.19 to 0.41)<br>[-0.25 to 0.47] | 0.8705<br>(0.67 to 1.07)*<br>[0.63 to 1.11]* | 0.3993<br>(0.12 to 0.68)*<br>[0.06 to 0.74]* | 0.3887<br>(0.17 to 0.61)*<br>[0.13 to 0.65]* | 0.4883<br>(0.30 to 0.68)*<br>[0.26 to 0.72]* | 5 | 5 |
| **4) Marriage and Sexual Activity** | | | | | | | | |
| Mother got married younger than 18 years old | -0.3415<br>(-0.60 to -0.08)*<br>[-0.65 to -0.03]* | -0.0744<br>(-0.40 to 0.25)<br>[-0.46 to 0.31] | -0.0690<br>(-0.24 to 0.10)<br>[-0.27 to 0.13] | -0.0211<br>(-0.31 to 0.26)<br>[-0.36 to 0.32] | 0.0933<br>(-0.14 to 0.32)<br>[-0.18 to 0.37] | 0.0458<br>(-0.16 to 0.25)<br>[-0.19 to 0.29] | 1 | 1 |
| Mother had first intercourse before 15 years old | -0.1735<br>(-0.42 to 0.07)<br>[-0.47 to 0.12] | -0.1824<br>(-0.60 to 0.24)<br>[-0.68 to 0.31] | -0.0461<br>(-0.25 to 0.15)<br>[-0.28 to 0.19] | -0.4738<br>(-0.82 to -0.13)*<br>[-0.88 to -0.06]* | 0.0348<br>(-0.21 to 0.28)<br>[-0.26 to 0.33] | -0.2409<br>(-0.46 to -0.02)*<br>[-0.51 to 0.03] | 2 | 1 |
| **5) Fertility** | | | | | | | | |
| Mother had first child before 18 years old | 0.3807<br>(-0.12 to 0.88)<br>[-0.22 to 0.98] | -0.2632<br>(-0.95 to 0.42)<br>[-1.08 to 0.55] | 0.2788<br>(-0.09 to 0.65)<br>[-0.16 to 0.72] | -0.1290<br>(-0.74 to 0.48)<br>[-0.86 to 0.60] | 0.1266<br>(-0.34 to 0.59)<br>[-0.43 to 0.68] | -0.1918<br>(-0.61 to 0.23)<br>[-0.69 to 0.31] | 0 | 0 |
| **6) Fertility Preferences** | | | | | | | | |
| Mother wanted pregnancy (wanted before or during pregnancy) | -0.3603<br>(-0.91 to 0.19)<br>[-1.02 to 0.30] | 0.4128<br>(-0.55 to 1.38)<br>[-0.74 to 1.56] | 1.1169<br>(0.48 to 1.76)*<br>[0.35 to 1.88]* | 2.1399<br>(0.71 to 3.57)*<br>[0.44 to 3.84]* | -0.0408<br>(-0.55 to 0.47)<br>[-0.65 to 0.56] | -0.5186<br>(-1.36 to 0.33)<br>[-1.53 to 0.49] | 2 | 2 |
| **7) Family Planning** | | | | | | | | |
| Mother visited health facility in last 12 months | 0.8784<br>(0.67 to 1.09)*<br>[0.63 to 1.12]* | 0.5858<br>(0.27 to 0.90)*<br>[0.21 to 0.96]* | 0.3664<br>(0.20 to 0.53)*<br>[0.17 to 0.56]* | 0.8284<br>(0.55 to 1.10)*<br>[0.50 to 1.16]* | 0.5999<br>(0.36 to 0.84)*<br>[0.31 to 0.89]* | 0.5543<br>(0.36 to 0.75)*<br>[0.33 to 0.78]* | 6 | 6 |
| Mother uses modern contraceptive method | 1.9052<br>(1.16 to 2.65)*<br>[1.01 to 2.80]* | 1.3205<br>(0.44 to 2.20)*<br>[0.27 to 2.37]* | 0.2820<br>(-0.41 to 0.98)<br>[-0.55 to 1.11] | 2.7720<br>(1.87 to 3.67)*<br>[1.70 to 3.84]* | 0.9733<br>(0.23 to 1.72)*<br>[0.09 to 1.86]* | 1.6639<br>(1.12 to 2.21)*<br>[1.02 to 2.31]* | 5 | 5 |
| Mother has used or plans to use contraceptive methods | 2.1138<br>(1.46 to 2.76)*<br>[1.34 to 2.89]* | 0.4297<br>(-0.86 to 1.72)<br>[-1.11 to 1.97] | 0.4381<br>(0.13 to 0.74)*<br>[0.08 to 0.80]* | 0.9479<br>(0.45 to 1.45)*<br>[0.36 to 1.54]* | 1.5829<br>(0.87 to 2.29)*<br>[0.74 to 2.43]* | 1.7129<br>(1.13 to 2.30)*<br>[1.02 to 2.41]* | 5 | 5 |
| Mother participates in family planning decisions | 1.4832<br>(0.99 to 1.98)*<br>[0.89 to 2.08]* | -0.3622<br>(-1.02 to 0.30)<br>[-1.15 to 0.42] | 0.1734<br>(-0.32 to 0.66)<br>[-0.41 to 0.76] | 0.6685<br>(0.16 to 1.18)*<br>[0.06 to 1.27]* | 0.1280<br>(-0.33 to 0.59)<br>[-0.42 to 0.67] | 1.2722<br>(0.81 to 1.74)*<br>[0.72 to 1.83]* | 3 | 3 |
| Mother heard about family planning in the radio (last months) | 0.9516<br>(0.72 to 1.18)*<br>[0.68 to 1.22]* | 0.3918<br>(0.06 to 0.72)*<br>[0.00 to 0.79] | 0.2673<br>(0.12 to 0.41)*<br>[0.09 to 0.44]* | -0.1027<br>(-0.34 to 0.14)<br>[-0.39 to 0.18] | 0.3675<br>(0.13 to 0.61)*<br>[0.08 to 0.65]* | 0.3568<br>(0.11 to 0.61)*<br>[0.06 to 0.66]* | 5 | 4 |
| Mother heard about family planning in TV (last months) | 0.5267<br>(0.15 to 0.90)*<br>[0.08 to 0.97]* | 0.1394<br>(-0.25 to 0.53)<br>[-0.33 to 0.61] | 0.3480<br>(0.18 to 0.52)*<br>[0.14 to 0.55]* | -0.3971<br>(-0.66 to -0.13)*<br>[-0.72 to -0.08]* | 0.3929<br>(0.00 to 0.79)<br>[-0.08 to 0.86] | 0.1903<br>(-0.16 to 0.54)<br>[-0.22 to 0.60] | 3 | 3 |
| Mother read about family planning in newspaper (last months) | 1.1192<br>(0.40 to 1.84)*<br>[0.26 to 1.98]* | 1.8025<br>(0.78 to 2.83)*<br>[0.58 to 3.02]* | 0.6646<br>(0.26 to 1.07)*<br>[0.19 to 1.14]* | -0.8186<br>(-1.26 to -0.38)*<br>[-1.34 to -0.30]* | 0.0401<br>(-0.35 to 0.43)<br>[-0.42 to 0.50] | 0.1793<br>(-0.33 to 0.69)<br>[-0.43 to 0.79] | 4 | 4 |
| Mother was told side- | 1.6451 | 1.9893 | 0.5847 | 1.4773 | 1.3570 | 0.4071 | 5 | 4 |

| | | | | | | | | |
|---|---|---|---|---|---|---|---|---|
| effects when getting contraceptives | (0.92 to 2.37)*<br>[0.78 to 2.51]* | (1.16 to 2.82)*<br>[1.00 to 2.98]* | (-0.03 to 1.20)<br>[-0.15 to 1.32] | (0.86 to 2.09)*<br>[0.75 to 2.21]* | (0.68 to 2.03)*<br>[0.55 to 2.16]* | (0.05 to 0.77)*<br>[-0.02 to 0.84] | | |
| Mother's number of known contraceptive methods (out of 9) | 0.7554<br>(0.62 to 0.89)*<br>[0.60 to 0.91]* | 0.4353<br>(0.26 to 0.62)*<br>[0.22 to 0.65]* | 0.4185<br>(0.29 to 0.55)*<br>[0.26 to 0.58]* | 0.3951<br>(0.22 to 0.57)*<br>[0.18 to 0.61]* | 0.5183<br>(0.34 to 0.70)*<br>[0.31 to 0.73]* | 0.3064<br>(0.19 to 0.43)*<br>[0.16 to 0.45]* | 6 | 6 |
| Mothers knows her ovulatory cycle | 0.2595<br>(0.05 to 0.47)*<br>[0.01 to 0.51]* | -0.0266<br>(-0.66 to 0.61)<br>[-0.78 to 0.73] | 0.0648<br>(-0.10 to 0.23)<br>[-0.13 to 0.26] | 0.7357<br>(0.48 to 0.99)*<br>[0.43 to 1.04]* | 0.2479<br>(0.00 to 0.50)<br>[-0.05 to 0.55] | 0.3852<br>(0.13 to 0.64)*<br>[0.09 to 0.68]* | 3 | 3 |
| **9) Reproductive Health** | | | | | | | | |
| Mother had at least 1 antenatal visit during pregnancy | 1.1325<br>(0.91 to 1.36)*<br>[0.87 to 1.40]* | 0.0585<br>(-0.42 to 0.54)<br>[-0.51 to 0.63] | 0.7641<br>(0.55 to 0.98)*<br>[0.51 to 1.02]* | 0.2276<br>(-0.23 to 0.69)<br>[-0.32 to 0.78] | 0.5445<br>(0.21 to 0.88)*<br>[0.15 to 0.94]* | 0.5325<br>(0.15 to 0.91)*<br>[0.08 to 0.99]* | 4 | 4 |
| Mother had blood pressure taken during pregnancy | 0.9983<br>(0.73 to 1.27)*<br>[0.67 to 1.32]* | 0.3391<br>(0.00 to 0.68)<br>[-0.07 to 0.74] | 0.9619<br>(0.74 to 1.18)*<br>[0.70 to 1.22]* | 0.2511<br>(-0.22 to 0.72)<br>[-0.31 to 0.81] | 0.3924<br>(0.15 to 0.63)*<br>[0.11 to 0.68]* | 0.3663<br>(0.07 to 0.67)*<br>[0.01 to 0.72]* | 4 | 4 |
| Mother had urine sample taken during pregnancy | 0.7070<br>(0.32 to 1.09)*<br>[0.25 to 1.17]* | 0.0461<br>(-0.24 to 0.33)<br>[-0.30 to 0.39] | 0.3315<br>(0.16 to 0.50)*<br>[0.13 to 0.54]* | 0.3998<br>(0.03 to 0.77)*<br>[-0.04 to 0.84] | 0.0761<br>(-0.19 to 0.34)<br>[-0.24 to 0.39] | 0.1959<br>(0.01 to 0.38)*<br>[-0.02 to 0.41] | 4 | 2 |
| Mother had blood sample taken during pregnancy | 0.8511<br>(0.45 to 1.25)*<br>[0.37 to 1.33]* | 0.1844<br>(-0.10 to 0.47)<br>[-0.15 to 0.52] | 0.1403<br>(-0.01 to 0.29)<br>[-0.04 to 0.32] | 0.4323<br>(0.15 to 0.72)*<br>[0.09 to 0.77]* | 0.2220<br>(-0.01 to 0.46)<br>[-0.06 to 0.50] | 0.5196<br>(0.22 to 0.82)*<br>[0.16 to 0.88]* | 3 | 3 |
| Mother had blood or urine sample taken during pregnancy | 0.7348<br>(0.37 to 1.09)*<br>[0.31 to 1.16]* | 0.0887<br>(-0.21 to 0.39)<br>[-0.27 to 0.44] | 0.3479<br>(0.17 to 0.52)*<br>[0.14 to 0.56]* | 0.4258<br>(0.01 to 0.84)*<br>[-0.07 to 0.92] | 0.1870<br>(-0.04 to 0.42)<br>[-0.09 to 0.46] | 0.6631<br>(0.35 to 0.98)*<br>[0.29 to 1.03]* | 4 | 3 |
| Mother assisted during delivery by skilled provider | 0.7062<br>(0.28 to 1.14)*<br>[0.19 to 1.22]* | 0.6246<br>(0.34 to 0.91)*<br>[0.29 to 0.96]* | 0.0015<br>(-0.18 to 0.18)<br>[-0.22 to 0.22] | 0.4033<br>(0.11 to 0.69)*<br>[0.06 to 0.75]* | 0.1491<br>(-0.09 to 0.39)<br>[-0.14 to 0.44] | 0.4695<br>(0.25 to 0.69)*<br>[0.21 to 0.73]* | 4 | 4 |
| Mother delivered at home | -0.6224<br>(-1.03 to -0.21)*<br>[-1.11 to -0.13]* | -0.6888<br>(-0.97 to -0.41)*<br>[-1.03 to -0.35]* | -0.2565<br>(-0.44 to -0.08)*<br>[-0.47 to -0.04]* | -0.6819<br>(-1.00 to -0.37)*<br>[-1.06 to -0.31]* | -0.0393<br>(-0.27 to 0.20)<br>[-0.32 to 0.24] | -0.6109<br>(-0.85 to -0.37)*<br>[-0.90 to -0.32]* | 5 | 5 |
| Mother says money is a problem to access healthcare when sick | -0.5799<br>(-0.83 to -0.33)*<br>[-0.88 to -0.28]* | -0.7088<br>(-1.02 to -0.40)*<br>[-1.08 to -0.33]* | 0.0132<br>(-0.14 to 0.17)<br>[-0.17 to 0.19] | 0.1055<br>(-0.15 to 0.36)<br>[-0.19 to 0.40] | -0.2951<br>(-0.53 to -0.06)*<br>[-0.57 to -0.02]* | -0.0933<br>(-0.29 to 0.11)<br>[-0.33 to 0.14] | 3 | 3 |
| Mother says distance is a problem to access healthcare when sick | -0.5541<br>(-0.77 to -0.34)*<br>[-0.81 to -0.30]* | -0.4695<br>(-0.77 to -0.17)*<br>[-0.83 to -0.11]* | 0.0116<br>(-0.15 to 0.17)<br>[-0.18 to 0.20] | -0.0646<br>(-0.34 to 0.21)<br>[-0.39 to 0.26] | -0.1376<br>(-0.35 to 0.08)<br>[-0.39 to 0.12] | -0.1261<br>(-0.32 to 0.07)<br>[-0.36 to 0.11] | 2 | 2 |
| Mother says getting permission to go is a problem to access healthcare when sick | -0.3399<br>(-0.58 to -0.10)*<br>[-0.62 to -0.06]* | -0.3798<br>(-0.65 to -0.11)*<br>[-0.71 to -0.05]* | 0.0141<br>(-0.29 to 0.32)<br>[-0.35 to 0.38] | -0.2822<br>(-0.65 to 0.08)<br>[-0.72 to 0.15] | -0.2767<br>(-0.77 to 0.22)<br>[-0.86 to 0.31] | -0.6095<br>(-0.99 to -0.23)*<br>[-1.06 to -0.16]* | 3 | 3 |
| **10) Child health** | | | | | | | | |
| Mother knows Oral Rehydration Salts packets | 1.3809<br>(0.84 to 1.92)*<br>[0.74 to 2.02]* | 1.7369<br>(0.51 to 2.96)*<br>[0.28 to 3.20]* | 0.4087<br>(0.25 to 0.56)*<br>[0.22 to 0.59]* | 0.4542<br>(0.20 to 0.71)*<br>[0.15 to 0.76]* | 0.3743<br>(-0.10 to 0.85)<br>[-0.20 to 0.94] | 0.9903<br>(0.44 to 1.54)*<br>[0.34 to 1.64]* | 5 | 5 |
| **11) Nutrition of Children & Adults** | | | | | | | | |
| Mother ever breastfed her child | 0.2008<br>(-1.84 to 2.24)<br>[-2.23 to 2.63] | -7.6832<br>(-54.3 to 38.9)<br>[-63.2 to 47.8] | 0.0910<br>(-0.55 to 0.74)<br>[-0.68 to 0.86] | 1.7803<br>(0.70 to 2.86)*<br>[0.50 to 3.07]* | -6.0166<br>(-49.1 to 37.0)<br>[-57.3 to 45.3] | 1.3364<br>(0.63 to 2.05)*<br>[0.49 to 2.18]* | 2 | 2 |
| Mother breastfed her child the day it was born | 0.4715<br>(0.24 to 0.70)*<br>[0.20 to 0.74]* | 0.5992<br>(0.25 to 0.95)*<br>[0.18 to 1.02]* | 0.5716<br>(0.40 to 0.74)*<br>[0.37 to 0.78]* | 0.1960<br>(-0.17 to 0.56)<br>[-0.24 to 0.63] | -0.0191<br>(-0.37 to 0.33)<br>[-0.44 to 0.40] | 0.5120<br>(0.18 to 0.84)*<br>[0.12 to 0.91]* | 4 | 4 |
| **15) Women's Empowerment** | | | | | | | | |
| Mother justifies being beaten for going out without permission | -0.2061<br>(-0.49 to 0.08)<br>[-0.55 to 0.14] | -0.1819<br>(-0.56 to 0.20)<br>[-0.64 to 0.27] | 0.0590<br>(-0.09 to 0.21)<br>[-0.12 to 0.24] | -0.1681<br>(-0.42 to 0.08)<br>[-0.47 to 0.13] | 0.2536<br>(-0.01 to 0.52)<br>[-0.06 to 0.57] | -0.1845<br>(-0.38 to 0.01)<br>[-0.41 to 0.04] | 0 | 0 |
| Mother justifies being | -0.2076 | -0.0286 | -0.0198 | -0.1559 | -0.0788 | 0.0258 | 0 | 0 |

| | | | | | | | | |
|---|---|---|---|---|---|---|---|---|
| beaten for neglecting a child | (-0.44 to 0.02)<br>[-0.48 to 0.06] | (-0.33 to 0.27)<br>[-0.38 to 0.33] | (-0.17 to 0.13)<br>[-0.20 to 0.16] | (-0.41 to 0.10)<br>[-0.46 to 0.14] | (-0.29 to 0.14)<br>[-0.33 to 0.18] | (-0.16 to 0.21)<br>[-0.20 to 0.25] | | |
| Mother justifies being beaten for arguing with partner | -0.4812<br>(-0.82 to -0.15)*<br>[-0.88 to -0.08]* | -0.0672<br>(-0.49 to 0.36)<br>[-0.57 to 0.44] | 0.0828<br>(-0.07 to 0.23)<br>[-0.09 to 0.26] | 0.0648<br>(-0.19 to 0.32)<br>[-0.24 to 0.37] | -0.1403<br>(-0.35 to 0.07)<br>[-0.39 to 0.11] | -0.1171<br>(-0.31 to 0.07)<br>[-0.34 to 0.11] | 1 | 1 |
| Mother justifies being beaten for not having sex | -0.3615<br>(-0.88 to 0.16)<br>[-0.98 to 0.26] | -0.5852<br>(-1.17 to 0.00)*<br>[-1.28 to 0.11] | 0.0568<br>(-0.09 to 0.20)<br>[-0.12 to 0.23] | -0.1180<br>(-0.37 to 0.14)<br>[-0.42 to 0.18] | -0.1475<br>(-0.35 to 0.06)<br>[-0.39 to 0.10] | -0.2078<br>(-0.40 to -0.02)*<br>[-0.43 to 0.02] | 2 | 0 |
| Mother justifies being beaten for burning food | -0.5470<br>(-0.94 to -0.15)*<br>[-1.01 to -0.08]* | -0.9143<br>(-1.45 to -0.38)*<br>[-1.56 to -0.27]* | -0.1559<br>(-0.32 to 0.01)<br>[-0.35 to 0.04] | -0.0253<br>(-0.30 to 0.25)<br>[-0.35 to 0.30] | -0.1625<br>(-0.37 to 0.04)<br>[-0.41 to 0.08] | -0.0147<br>(-0.23 to 0.20)<br>[-0.27 to 0.24] | 2 | 2 |
| Mother justifies being beaten | -0.1713<br>(-0.39 to 0.05)<br>[-0.43 to 0.09] | -0.0827<br>(-0.36 to 0.20)<br>[-0.42 to 0.25] | 0.0810<br>(-0.08 to 0.24)<br>[-0.11 to 0.27] | 0.0439<br>(-0.20 to 0.29)<br>[-0.25 to 0.34] | 0.3404<br>(0.03 to 0.66)*<br>[-0.03 to 0.72] | -0.1711<br>(-0.35 to 0.01)<br>[-0.38 to 0.04] | 1 | 0 |
| Mother decides on her own health | 0.3160<br>(0.07 to 0.56)*<br>[0.02 to 0.61]* | 0.2455<br>(-0.04 to 0.53)<br>[-0.09 to 0.58] | 0.2242<br>(0.02 to 0.42)*<br>[-0.01 to 0.46] | 0.0222<br>(-0.34 to 0.38)<br>[-0.41 to 0.45] | -0.2602<br>(-0.49 to -0.03)*<br>[-0.53 to 0.01] | 0.5016<br>(0.27 to 0.73)*<br>[0.23 to 0.78]* | 4 | 2 |
| Mother decides on large household purchases | 0.0936<br>(-0.15 to 0.34)<br>[-0.20 to 0.38] | -0.1231<br>(-0.43 to 0.18)<br>[-0.49 to 0.24] | -0.1087<br>(-0.31 to 0.09)<br>[-0.35 to 0.13] | 0.0613<br>(-0.31 to 0.43)<br>[-0.38 to 0.50] | 0.0419<br>(-0.20 to 0.28)<br>[-0.24 to 0.33] | 0.1260<br>(-0.08 to 0.33)<br>[-0.12 to 0.37] | 0 | 0 |
| Mother decides on visits to family or relatives | 0.2169<br>(-0.01 to 0.45)<br>[-0.06 to 0.49] | -0.0817<br>(-0.37 to 0.21)<br>[-0.43 to 0.26] | -0.0936<br>(-0.25 to 0.07)<br>[-0.28 to 0.10] | -0.1924<br>(-0.53 to 0.15)<br>[-0.60 to 0.21] | 0.0788<br>(-0.15 to 0.31)<br>[-0.20 to 0.35] | 0.3332<br>(0.12 to 0.55)*<br>[0.08 to 0.59]* | 1 | 1 |

*Data sources: Various DHS [1].*
*Coefficients were obtained from ordinal logistic regressions controlling for the variables in Table 4; children groups LV, MV, and 8V were used as ordinal categories.*
** denotes CIs not containing zero. Values represent median of the average value of the coefficient, parentheses "()" show 90% CI, brackets "[]" show 95% CI.*

*Table 7: Average values of tested variables, by DHS dataset and vaccine group*

| Variables | Vaccine group | Nepal | | Senegal | | Zambia | | Count of differences/ coefficients not containing zero (95% CI) | |
|---|---|---|---|---|---|---|---|---|---|
| | | 2001† | 2016‡ | 2005† | 2019‡ | 2001-02† | 2018‡ | 8V-LV difference | OLR coefficient |
| **3) Respondent's characteristics** | | | | | | | | | |
| Mother's years of education | LV | 0.2 (0.0 to 0.4) | 2.7 (1.7 to 3.9) | 0.3 (0.1 to 0.4) | 1.1 (0.6 to 1.7) | 4.1 (3.2 to 4.9) | 4.5 (3.8 to 5.3) | | |
| | MV | 0.3 (0.1 to 0.7) | 3.0 (1.7 to 4.4) | 0.6 (0.3 to 1.0) | 1.6 (0.9 to 2.6) | 4.2 (3.2 to 5.2) | 5.6 (4.7 to 6.4) | | |
| | 8V | 0.9 (0.4 to 1.5) | 3.5 (2.2 to 4.9) | 1.4 (0.9 to 1.9) | 1.7 (1.0 to 2.7) | 4.9 (4.0 to 5.8) | 6.0 (5.2 to 6.9) | | |
| | 8V-LV | 0.7 (0.2 to 1.3)* | 0.8 (-0.6 to 2.1) | 1.1 (0.6 to 1.7)* | 0.6 (-0.3 to 1.6) | 0.8 (-0.2 to 1.8) | 1.5 (0.6 to 2.4)* | 3 | 4 |
| Mother is literate | LV | 3% (0% to 8%) | 36% (24% to 52%) | 3% (1% to 7%) | 14% (6% to 22%) | 42% (30% to 54%) | 33% (23% to 44%) | | |
| | MV | 14% (8% to 23%) | 50% (31% to 67%) | 11% (6% to 17%) | 23% (13% to 35%) | 42% (29% to 55%) | 47% (36% to 60%) | | |
| | 8V | 26% (17% to 36%) | 52% (36% to 69%) | 21% (14% to 28%) | 23% (14% to 35%) | 54% (41% to 67%) | 53% (41% to 66%) | | |
| | 8V-LV | 22% (12% to 32%)* | 14% (-7% to 36%) | 17% (10% to 26%)* | 10% (-3% to 22%) | 12% (-4% to 28%) | 20% (5% to 36%)* | 3 | 5 |
| **4) Marriage and Sexual Activity** | | | | | | | | | |
| Mother got married younger than 18 years old | LV | 71% (61% to 80%) | 67% (50% to 79%) | 67% (58% to 74%) | 51% (40% to 62%) | 57% (45% to 68%) | 48% (38% to 59%) | | |
| | MV | 73% (63% to 83%) | 62% (45% to 79%) | 67% (58% to 76%) | 56% (44% to 68%) | 57% (43% to 70%) | 44% (32% to 57%) | | |
| | 8V | 69% (57% to 79%) | 62% (45% to 79%) | 68% (59% to 76%) | 53% (39% to 65%) | 59% (45% to 72%) | 44% (32% to 56%) | | |
| | 8V-LV | -2% (-16% to 11%) | -5% (-24% to 14%) | 1% (-10% to 13%) | 3% (-13% to 17%) | 3% (-13% to 17%) | -5% (-18% to 11%) | 0 | 1 |
| Mother had first intercourse before 15 years old | LV | 28% (20% to 38%) | 19% (7% to 29%) | 27% (20% to 35%) | 19% (12% to 30%) | 26% (17% to 38%) | 32% (23% to 43%) | | |
| | MV | 23% (14% to 33%) | 12% (2% to 24%) | 22% (15% to 31%) | 25% (16% to 35%) | 23% (13% to 36%) | 24% (15% to 34%) | | |
| | 8V | 23% (14% to 33%) | 12% (2% to 21%) | 23% (16% to 31%) | 17% (9% to 29%) | 23% (13% to 36%) | 21% (11% to 31%) | | |
| | 8V-LV | -6% (-18% to 7%) | -7% (-21% to 7%) | -4% (-14% to 6%) | -3% (-14% to 9%) | -3% (-19% to 13%) | -11% (-25% to 2%) | 0 | 1 |
| **5) Fertility** | | | | | | | | | |
| Mother had first child before 18 years old | LV | 28% (19% to 37%) | 31% (17% to 45%) | 49% (41% to 58%) | 26% (17% to 36%) | 48% (36% to 59%) | 46% (36% to 55%) | | |
| | MV | 28% (17% to 39%) | 24% (10% to 40%) | 44% (35% to 54%) | 39% (27% to 52%) | 41% (28% to 55%) | 46% (34% to 58%) | | |
| | 8V | 36% (24% to 48%) | 24% (10% to 38%) | 49% (39% to 58%) | 34% (21% to 47%) | 46% (32% to 61%) | 41% (30% to 54%) | | |
| | 8V-LV | 9% (-6% to 21%) | -7% (-26% to 12%) | 0% (-12% to 10%) | 6% (-6% to 21%) | -3% (-19% to 14%) | -3% (-17% to 10%) | 0 | 0 |
| **6) Fertility Preferences** | | | | | | | | | |

| | | | | | | | | | |
|---|---|---|---|---|---|---|---|---|---|
| Mother wanted pregnancy (wanted before or during pregnancy) | LV | 73%<br>(63% to 82%) | 81%<br>(69% to 91%) | 92%<br>(88% to 97%) | 99%<br>(96% to 100%) | 75%<br>(65% to 84%) | 94%<br>(90% to 99%) | | |
| | MV | 66%<br>(54% to 77%) | 93%<br>(81% to 100%) | 93%<br>(88% to 97%) | 99%<br>(91% to 100%) | 77%<br>(65% to 87%) | 95%<br>(90% to 100%) | | |
| | 8V | 62%<br>(52% to 73%) | 90%<br>(76% to 98%) | 95%<br>(90% to 98%) | 100%<br>(96% to 100%) | 74%<br>(62% to 86%) | 94%<br>(88% to 99%) | | |
| | 8V-LV | -10%<br>(-22% to 2%) | 10%<br>(-7% to 24%) | 3%<br>(-3% to 8%) | 0%<br>(-3% to 4%) | 0%<br>(-16% to 14%) | -1%<br>(-7% to 6%) | 0 | 2 |
| **7) Family Planning** | | | | | | | | | |
| Mother visited health facility in last 12 months | LV | 36%<br>(26% to 46%) | 69%<br>(55% to 81%) | 55%<br>(47% to 64%) | 64%<br>(53% to 74%) | 59%<br>(46% to 71%) | 54%<br>(44% to 64%) | | |
| | MV | 50%<br>(39% to 61%) | 71%<br>(55% to 86%) | 65%<br>(56% to 74%) | 79%<br>(69% to 88%) | 75%<br>(64% to 86%) | 71%<br>(61% to 82%) | | |
| | 8V | 66%<br>(53% to 76%) | 83%<br>(69% to 95%) | 70%<br>(62% to 78%) | 81%<br>(70% to 90%) | 83%<br>(71% to 91%) | 76%<br>(66% to 85%) | | |
| | 8V-LV | 30%<br>(16% to 44%)* | 14%<br>(-5% to 33%) | 15%<br>(3% to 26%)* | 17%<br>(1% to 31%)* | 23%<br>(9% to 38%)* | 22%<br>(7% to 36%)* | 5 | 6 |
| Mother uses modern contraceptive method | LV | 14%<br>(8% to 22%) | 24%<br>(12% to 38%) | 5%<br>(2% to 9%) | 9%<br>(4% to 16%) | 16%<br>(9% to 26%) | 26%<br>(18% to 37%) | | |
| | MV | 20%<br>(11% to 29%) | 31%<br>(17% to 48%) | 8%<br>(3% to 14%) | 17%<br>(8% to 27%) | 25%<br>(13% to 38%) | 51%<br>(39% to 61%) | | |
| | 8V | 29%<br>(18% to 39%) | 38%<br>(24% to 57%) | 8%<br>(4% to 14%) | 25%<br>(16% to 36%) | 26%<br>(14% to 39%) | 57%<br>(45% to 68%) | | |
| | 8V-LV | 13%<br>(1% to 27%)* | 14%<br>(-5% to 36%) | 3%<br>(-2% to 9%) | 17%<br>(5% to 29%)* | 10%<br>(-4% to 23%) | 30%<br>(16% to 45%)* | 3 | 5 |
| Mother has used or plans to use contraceptive methods | LV | 79%<br>(71% to 87%) | 100%<br>(100% to 100%) | 35%<br>(27% to 43%) | 34%<br>(22% to 45%) | 84%<br>(74% to 91%) | 78%<br>(69% to 86%) | | |
| | MV | 87%<br>(78% to 93%) | 93%<br>(81% to 100%) | 43%<br>(33% to 51%) | 52%<br>(39% to 64%) | 91%<br>(84% to 99%) | 86%<br>(78% to 94%) | | |
| | 8V | 92%<br>(86% to 98%) | 95%<br>(88% to 100%) | 44%<br>(36% to 53%) | 57%<br>(45% to 70%) | 94%<br>(88% to 100%) | 93%<br>(87% to 98%) | | |
| | 8V-LV | 13%<br>(3% to 23%)* | -5%<br>(-12% to 0%)* | 10%<br>(-1% to 21%) | 23%<br>(6% to 40%)* | 12%<br>(1% to 22%)* | 15%<br>(6% to 26%)* | 5 | 5 |
| Mother participates in family planning decisions | LV | 14%<br>(8% to 22%) | 81%<br>(67% to 91%) | 3%<br>(1% to 7%) | 48%<br>(38% to 60%) | 30%<br>(20% to 42%) | 45%<br>(33% to 55%) | | |
| | MV | 21%<br>(13% to 30%) | 81%<br>(67% to 93%) | 8%<br>(3% to 13%) | 56%<br>(43% to 68%) | 35%<br>(23% to 46%) | 61%<br>(48% to 74%) | | |
| | 8V | 30%<br>(20% to 41%) | 76%<br>(62% to 90%) | 6%<br>(3% to 12%) | 64%<br>(52% to 75%) | 35%<br>(23% to 49%) | 63%<br>(51% to 76%) | | |
| | 8V-LV | 14%<br>(2% to 28%)* | -2%<br>(-21% to 17%) | 3%<br>(-2% to 8%) | 16%<br>(0% to 31%)* | 4%<br>(-10% to 20%) | 20%<br>(5% to 32%)* | 3 | 3 |
| Mother heard about family planning in the radio (last months) | LV | 29%<br>(20% to 39%) | 2%<br>(0% to 10%) | 36%<br>(29% to 43%) | 39%<br>(29% to 51%) | 41%<br>(29% to 51%) | 9%<br>(5% to 16%) | | |
| | MV | 42%<br>(31% to 54%) | 19%<br>(7% to 31%) | 41%<br>(33% to 50%) | 49%<br>(38% to 62%) | 29%<br>(16% to 42%) | 13%<br>(6% to 22%) | | |
| | 8V | 54%<br>(43% to 67%) | 26%<br>(12% to 43%) | 49%<br>(40% to 58%) | 43%<br>(31% to 56%) | 41%<br>(28% to 54%) | 15%<br>(8% to 24%) | | |
| | 8V-LV | 26%<br>(11% to 40%)* | 24%<br>(9% to 40%)* | 13%<br>(1% to 24%)* | 4%<br>(-10% to 20%) | 0%<br>(-13% to 16%) | 6%<br>(-3% to 16%) | 3 | 4 |
| Mother heard about family planning in TV (last | LV | 2%<br>(0% to 6%) | 10%<br>(2% to 19%) | 15%<br>(10% to 21%) | 34%<br>(25% to 44%) | 17%<br>(9% to 28%) | 5%<br>(1% to 9%) | 1 | 3 |
| | MV | 6%<br>(1% to 11%) | 14%<br>(2% to 29%) | 20%<br>(13% to 28%) | 40%<br>(29% to 53%) | 13%<br>(3% to 23%) | 6%<br>(1% to 13%) | | |

| | | | | | | | | | |
|---|---|---|---|---|---|---|---|---|---|
| months) | 8V | 8%<br>(2% to 14%) | 14%<br>(5% to 26%) | 24%<br>(16% to 31%) | 31%<br>(21% to 44%) | 16%<br>(7% to 28%) | 7%<br>(2% to 14%) | | |
| | 8V-LV | 6%<br>(0% to 11%)* | 5%<br>(-7% to 19%) | 8%<br>(-1% to 17%) | -3%<br>(-16% to 10%) | -1%<br>(-12% to 10%) | 2%<br>(-3% to 9%) | | |
| Mother read about family planning in newspaper (last months) | LV | 0%<br>(0% to 0%) | 0%<br>(0% to 0%) | 3%<br>(1% to 6%) | 0%<br>(0% to 0%) | 9%<br>(3% to 16%) | 0%<br>(0% to 0%) | | |
| | MV | 1%<br>(0% to 4%) | 0%<br>(0% to 2%) | 1%<br>(0% to 3%) | 5%<br>(1% to 12%) | 9%<br>(1% to 19%) | 2%<br>(0% to 7%) | | |
| | 8V | 2%<br>(0% to 7%) | 5%<br>(0% to 14%) | 2%<br>(0% to 5%) | 3%<br>(0% to 8%) | 9%<br>(1% to 16%) | 2%<br>(0% to 7%) | | |
| | 8V-LV | 2%<br>(0% to 7%)* | 5%<br>(0% to 14%)* | -1%<br>(-5% to 3%) | 3%<br>(0% to 8%)* | -1%<br>(-10% to 9%) | 2%<br>(0% to 7%)* | 4 | 4 |
| Mother was told side-effects when getting contraceptives | LV | 6%<br>(1% to 11%) | 10%<br>(2% to 19%) | 1%<br>(0% to 3%) | 9%<br>(3% to 16%) | 7%<br>(3% to 14%) | 21%<br>(13% to 30%) | | |
| | MV | 7%<br>(2% to 14%) | 10%<br>(0% to 19%) | 5%<br>(1% to 9%) | 16%<br>(8% to 27%) | 10%<br>(3% to 17%) | 40%<br>(29% to 51%) | | |
| | 8V | 11%<br>(4% to 20%) | 24%<br>(12% to 40%) | 4%<br>(1% to 8%) | 21%<br>(13% to 32%) | 14%<br>(6% to 25%) | 41%<br>(30% to 53%) | | |
| | 8V-LV | 6%<br>(-3% to 16%) | 14%<br>(-2% to 33%) | 3%<br>(-1% to 7%) | 13%<br>(1% to 25%)* | 6%<br>(-3% to 17%) | 21%<br>(7% to 33%)* | 2 | 4 |
| Mother's number of known contraceptive methods (out of 9) | LV | 5.1<br>(4.6 to 5.5) | 6.9<br>(6.4 to 7.4) | 3.7<br>(3.3 to 4.1) | 4.9<br>(4.4 to 5.4) | 4.8<br>(4.3 to 5.4) | 5.7<br>(5.3 to 6.1) | | |
| | MV | 6.2<br>(5.8 to 6.6) | 7.5<br>(7.0 to 8.0) | 4.6<br>(4.2 to 5.0) | 5.4<br>(4.8 to 5.9) | 5.1<br>(4.6 to 5.5) | 6.6<br>(6.2 to 6.9) | | |
| | 8V | 6.6<br>(6.2 to 6.9) | 7.8<br>(7.3 to 8.2) | 4.9<br>(4.5 to 5.3) | 5.8<br>(5.2 to 6.3) | 5.6<br>(5.1 to 6.1) | 6.6<br>(6.3 to 7.0) | | |
| | 8V-LV | 1.5<br>(0.9 to 2.0)* | 0.9<br>(0.1 to 1.5)* | 1.1<br>(0.6 to 1.7)* | 0.9<br>(0.2 to 1.7)* | 0.7<br>(0.1 to 1.4)* | 0.9<br>(0.5 to 1.4)* | 6 | 6 |
| Mothers knows her ovulatory cycle | LV | 46%<br>(36% to 56%) | 93%<br>(86% to 100%) | 53%<br>(45% to 60%) | 52%<br>(42% to 64%) | 75%<br>(65% to 87%) | 76%<br>(67% to 84%) | | |
| | MV | 64%<br>(54% to 74%) | 98%<br>(90% to 100%) | 74%<br>(67% to 83%) | 61%<br>(49% to 73%) | 80%<br>(70% to 90%) | 89%<br>(79% to 95%) | | |
| | 8V | 64%<br>(54% to 76%) | 98%<br>(88% to 100%) | 70%<br>(62% to 78%) | 71%<br>(61% to 83%) | 84%<br>(74% to 93%) | 89%<br>(80% to 94%) | | |
| | 8V-LV | 20%<br>(4% to 33%)* | 2%<br>(-7% to 12%) | 17%<br>(6% to 28%)* | 19%<br>(5% to 34%)* | 8%<br>(-6% to 22%) | 13%<br>(0% to 23%)* | 4 | 3 |
| **9) Reproductive Health** | | | | | | | | | |
| Mother had at least 1 antenatal visit during pregnancy | LV | 7%<br>(2% to 13%) | 79%<br>(67% to 90%) | 63%<br>(55% to 70%) | 86%<br>(78% to 94%) | 77%<br>(65% to 87%) | 84%<br>(75% to 91%) | | |
| | MV | 26%<br>(17% to 36%) | 95%<br>(88% to 100%) | 83%<br>(76% to 90%) | 92%<br>(86% to 97%) | 87%<br>(78% to 94%) | 97%<br>(91% to 100%) | | |
| | 8V | 47%<br>(34% to 57%) | 90%<br>(79% to 98%) | 88%<br>(81% to 93%) | 94%<br>(86% to 99%) | 91%<br>(84% to 97%) | 95%<br>(91% to 99%) | | |
| | 8V-LV | 39%<br>(26% to 50%)* | 12%<br>(-7% to 26%) | 25%<br>(15% to 35%)* | 6%<br>(-3% to 17%) | 14%<br>(1% to 28%)* | 13%<br>(3% to 22%)* | 4 | 4 |
| Mother had blood pressure taken during pregnancy | LV | 6%<br>(2% to 11%) | 67%<br>(52% to 81%) | 56%<br>(48% to 64%) | 86%<br>(78% to 94%) | 61%<br>(48% to 72%) | 75%<br>(64% to 83%) | | |
| | MV | 12%<br>(6% to 20%) | 81%<br>(67% to 93%) | 82%<br>(76% to 89%) | 92%<br>(84% to 97%) | 72%<br>(61% to 84%) | 92%<br>(85% to 98%) | | |
| | 8V | 24%<br>(14% to 34%) | 83%<br>(69% to 95%) | 88%<br>(81% to 93%) | 94%<br>(86% to 99%) | 81%<br>(70% to 90%) | 91%<br>(84% to 97%) | | |
| | 8V-LV | 19%<br>(9% to 29%)* | 17%<br>(-2% to 36%) | 31%<br>(21% to 42%)* | 6%<br>(-1% to 17%) | 20%<br>(4% to 35%)* | 17%<br>(6% to 28%)* | 4 | 4 |

| | | | | | | | | | |
|---|---|---|---|---|---|---|---|---|---|
| Mother had urine sample taken during pregnancy | LV | 6% (1% to 10%) | 60% (45% to 74%) | 49% (40% to 57%) | 75% (66% to 84%) | 22% (13% to 32%) | 45% (34% to 55%) | | |
| | MV | 3% (0% to 8%) | 62% (45% to 79%) | 69% (61% to 77%) | 87% (78% to 94%) | 23% (13% to 35%) | 57% (45% to 69%) | | |
| | 8V | 8% (2% to 14%) | 64% (48% to 81%) | 72% (64% to 80%) | 87% (79% to 95%) | 20% (10% to 32%) | 60% (49% to 70%) | | |
| | 8V-LV | 2% (-4% to 9%) | 5% (-19% to 26%) | 24% (13% to 35%)* | 12% (0% to 25%) | -1% (-14% to 12%) | 14% (0% to 29%) | 1 | 2 |
| Mother had blood sample taken during pregnancy | LV | 4% (1% to 9%) | 50% (33% to 64%) | 30% (23% to 38%) | 60% (48% to 70%) | 25% (14% to 35%) | 76% (67% to 84%) | | |
| | MV | 2% (0% to 7%) | 52% (36% to 69%) | 47% (38% to 56%) | 66% (56% to 78%) | 33% (22% to 45%) | 92% (85% to 98%) | | |
| | 8V | 8% (2% to 14%) | 60% (40% to 76%) | 42% (33% to 50%) | 77% (65% to 87%) | 35% (23% to 48%) | 92% (86% to 98%) | | |
| | 8V-LV | 3% (-3% to 11%) | 10% (-12% to 31%) | 13% (0% to 24%)* | 17% (3% to 32%)* | 12% (-3% to 26%) | 16% (7% to 28%)* | 3 | 3 |
| Mother had blood or urine sample taken during pregnancy | LV | 6% (1% to 10%) | 62% (48% to 76%) | 49% (40% to 57%) | 81% (70% to 88%) | 29% (17% to 39%) | 76% (67% to 84%) | | |
| | MV | 4% (1% to 10%) | 67% (52% to 83%) | 72% (64% to 80%) | 88% (81% to 95%) | 33% (23% to 46%) | 93% (86% to 98%) | | |
| | 8V | 9% (3% to 17%) | 69% (50% to 83%) | 74% (65% to 81%) | 90% (82% to 96%) | 36% (25% to 49%) | 93% (87% to 99%) | | |
| | 8V-LV | 3% (-3% to 11%) | 7% (-17% to 26%) | 25% (14% to 36%)* | 10% (0% to 22%) | 9% (-6% to 23%) | 18% (8% to 29%)* | 2 | 3 |
| Mother assisted during delivery by skilled provider | LV | 6% (1% to 10%) | 43% (29% to 57%) | 30% (22% to 37%) | 51% (39% to 61%) | 28% (17% to 38%) | 56% (46% to 67%) | | |
| | MV | 2% (0% to 7%) | 45% (31% to 62%) | 38% (28% to 47%) | 58% (47% to 70%) | 35% (22% to 48%) | 75% (63% to 84%) | | |
| | 8V | 4% (1% to 10%) | 57% (40% to 74%) | 38% (28% to 47%) | 61% (48% to 73%) | 35% (22% to 46%) | 79% (69% to 87%) | | |
| | 8V-LV | 0% (-7% to 6%) | 14% (-7% to 33%) | 7% (-3% to 17%) | 10% (-3% to 25%) | 6% (-7% to 22%) | 22% (9% to 34%)* | 1 | 4 |
| Mother delivered at home | LV | 91% (86% to 97%) | 57% (43% to 74%) | 53% (46% to 61%) | 47% (36% to 58%) | 70% (58% to 80%) | 40% (29% to 49%) | | |
| | MV | 98% (93% to 100%) | 57% (40% to 71%) | 46% (37% to 56%) | 32% (22% to 44%) | 62% (49% to 75%) | 17% (10% to 28%) | | |
| | 8V | 93% (87% to 98%) | 40% (24% to 55%) | 42% (33% to 51%) | 29% (18% to 40%) | 64% (51% to 77%) | 15% (8% to 24%) | | |
| | 8V-LV | 2% (-7% to 9%) | -19% (-38% to 2%) | -12% (-23% to -2%)* | -18% (-32% to -5%)* | -6% (-20% to 9%) | -24% (-37% to -13%)* | 3 | 5 |
| Mother says money is a problem to access healthcare when sick | LV | 94% (89% to 99%) | 83% (71% to 95%) | 67% (60% to 75%) | 70% (60% to 79%) | 83% (74% to 91%) | 34% (25% to 45%) | | |
| | MV | 84% (76% to 91%) | 74% (62% to 90%) | 61% (52% to 69%) | 57% (44% to 69%) | 69% (57% to 81%) | 26% (16% to 37%) | | |
| | 8V | 81% (71% to 89%) | 69% (52% to 83%) | 65% (57% to 74%) | 65% (53% to 75%) | 70% (57% to 81%) | 26% (16% to 38%) | | |
| | 8V-LV | -13% (-24% to -4%)* | -14% (-33% to 5%) | -2% (-13% to 10%) | -5% (-21% to 9%) | -13% (-28% to 1%) | -7% (-22% to 6%) | 1 | 3 |
| Mother says distance is a problem to access healthcare when sick | LV | 80% (71% to 88%) | 79% (64% to 90%) | 45% (38% to 53%) | 48% (36% to 60%) | 65% (52% to 75%) | 52% (40% to 62%) | 1 | 2 |
| | MV | 67% (57% to 77%) | 71% (57% to 86%) | 46% (36% to 54%) | 42% (30% to 55%) | 59% (48% to 72%) | 39% (28% to 52%) | | |
| | 8V | 63% (50% to 73%) | 64% (48% to 81%) | 49% (40% to 58%) | 40% (29% to 52%) | 58% (43% to 70%) | 39% (29% to 51%) | | |

| | | | | | | | | | |
|---|---|---|---|---|---|---|---|---|---|
| | 8V-LV | -17%<br>(-29% to -4%)* | -14%<br>(-33% to 7%) | 3%<br>(-7% to 15%) | -8%<br>(-22% to 6%) | -6%<br>(-23% to 9%) | -13%<br>(-26% to 2%) | | |
| Mother says getting permission to go is a problem to access healthcare when sick | LV | 32%<br>(22% to 42%) | 45%<br>(29% to 60%) | 11%<br>(6% to 16%) | 16%<br>(8% to 23%) | 7%<br>(1% to 14%) | 15%<br>(8% to 23%) | | |
| | MV | 24%<br>(16% to 34%) | 38%<br>(21% to 55%) | 6%<br>(2% to 11%) | 21%<br>(12% to 31%) | 4%<br>(0% to 10%) | 5%<br>(0% to 10%) | | |
| | 8V | 22%<br>(13% to 32%) | 29%<br>(14% to 48%) | 8%<br>(3% to 13%) | 13%<br>(5% to 22%) | 4%<br>(0% to 10%) | 5%<br>(1% to 10%) | | |
| | 8V-LV | -10%<br>(-23% to 2%) | -17%<br>(-38% to 7%) | -3%<br>(-10% to 3%) | -3%<br>(-13% to 9%) | -3%<br>(-12% to 4%) | -10%<br>(-20% to -1%)* | 1 | 3 |
| **10) Child health** | | | | | | | | | |
| Mother knows Oral Rehydration Salts packets | LV | 91%<br>(84% to 96%) | 98%<br>(93% to 100%) | 50%<br>(42% to 58%) | 48%<br>(36% to 58%) | 90%<br>(83% to 97%) | 97%<br>(91% to 100%) | | |
| | MV | 94%<br>(88% to 99%) | 100%<br>(93% to 100%) | 58%<br>(49% to 66%) | 65%<br>(53% to 75%) | 96%<br>(91% to 100%) | 98%<br>(93% to 100%) | | |
| | 8V | 99%<br>(96% to 100%) | 100%<br>(98% to 100%) | 68%<br>(60% to 76%) | 69%<br>(56% to 79%) | 96%<br>(90% to 100%) | 99%<br>(95% to 100%) | | |
| | 8V-LV | 8%<br>(2% to 14%)* | 2%<br>(-2% to 7%) | 18%<br>(6% to 29%)* | 21%<br>(6% to 35%)* | 6%<br>(-3% to 13%) | 2%<br>(-2% to 8%) | 3 | 5 |
| **11) Nutrition of Children & Adults** | | | | | | | | | |
| Mother ever breastfed her child | LV | 100%<br>(100% to 100%) | 100%<br>(100% to 100%) | 94%<br>(90% to 97%) | 100%<br>(96% to 100%) | 100%<br>(100% to 100%) | 95%<br>(90% to 99%) | | |
| | MV | 100%<br>(100% to 100%) | 100%<br>(100% to 100%) | 99%<br>(97% to 100%) | 99%<br>(95% to 100%) | 100%<br>(100% to 100%) | 100%<br>(97% to 100%) | | |
| | 8V | 100%<br>(99% to 100%) | 100%<br>(95% to 100%) | 99%<br>(96% to 100%) | 100%<br>(99% to 100%) | 100%<br>(99% to 100%) | 100%<br>(97% to 100%) | | |
| | 8V-LV | 0%<br>(-1% to 0%)* | 0%<br>(-5% to 0%)* | 5%<br>(0% to 9%)* | 0%<br>(-1% to 3%) | 0%<br>(-1% to 0%)* | 3%<br>(0% to 9%)* | 5 | 2 |
| Mother breastfed her child the day it was born | LV | 78%<br>(69% to 86%) | 76%<br>(62% to 88%) | 67%<br>(59% to 74%) | 91%<br>(84% to 97%) | 88%<br>(81% to 96%) | 83%<br>(75% to 91%) | | |
| | MV | 69%<br>(58% to 79%) | 81%<br>(67% to 93%) | 74%<br>(66% to 82%) | 91%<br>(82% to 96%) | 93%<br>(84% to 99%) | 95%<br>(90% to 99%) | | |
| | 8V | 78%<br>(67% to 86%) | 88%<br>(76% to 98%) | 83%<br>(76% to 90%) | 92%<br>(84% to 97%) | 91%<br>(83% to 97%) | 94%<br>(88% to 99%) | | |
| | 8V-LV | 0%<br>(-12% to 11%) | 12%<br>(-5% to 29%) | 17%<br>(6% to 27%)* | 1%<br>(-8% to 9%) | 1%<br>(-9% to 12%) | 11%<br>(1% to 21%)* | 2 | 4 |
| **15) Women's Empowerment** | | | | | | | | | |
| Mother justifies being beaten for going out without permission | LV | 14%<br>(8% to 21%) | 24%<br>(10% to 36%) | 60%<br>(53% to 67%) | 51%<br>(39% to 61%) | 80%<br>(70% to 88%) | 34%<br>(25% to 45%) | | |
| | MV | 14%<br>(8% to 23%) | 12%<br>(2% to 24%) | 58%<br>(50% to 67%) | 48%<br>(36% to 62%) | 81%<br>(70% to 90%) | 38%<br>(26% to 49%) | | |
| | 8V | 10%<br>(4% to 18%) | 12%<br>(2% to 26%) | 60%<br>(52% to 69%) | 43%<br>(30% to 56%) | 84%<br>(74% to 93%) | 31%<br>(22% to 43%) | | |
| | 8V-LV | -4%<br>(-13% to 7%) | -10%<br>(-26% to 7%) | 1%<br>(-11% to 12%) | -6%<br>(-23% to 10%) | 4%<br>(-7% to 19%) | -3%<br>(-16% to 10%) | 0 | 0 |
| Mother justifies being beaten for neglecting a child | LV | 28%<br>(20% to 38%) | 24%<br>(12% to 38%) | 56%<br>(48% to 63%) | 47%<br>(35% to 58%) | 68%<br>(57% to 78%) | 39%<br>(30% to 51%) | | |
| | MV | 29%<br>(19% to 38%) | 29%<br>(14% to 45%) | 61%<br>(52% to 69%) | 51%<br>(38% to 64%) | 64%<br>(52% to 77%) | 37%<br>(26% to 48%) | | |
| | 8V | 21%<br>(12% to 30%) | 24%<br>(12% to 38%) | 59%<br>(50% to 68%) | 45%<br>(32% to 57%) | 67%<br>(54% to 78%) | 38%<br>(29% to 48%) | | |
| | 8V-LV | -7%<br>(-20% to 6%) | -2%<br>(-19% to 19%) | 3%<br>(-8% to 15%) | -3%<br>(-19% to 16%) | -1%<br>(-17% to 14%) | -1%<br>(-16% to 14%) | 0 | 0 |

| | | | | | | | | | |
|---|---|---|---|---|---|---|---|---|---|
| Mother justifies being beaten for arguing with partner | LV | 7% (2% to 12%) | 12% (2% to 24%) | 53% (45% to 61%) | 51% (39% to 62%) | 65% (54% to 75%) | 44% (33% to 55%) | | |
| | MV | 9% (3% to 17%) | 14% (5% to 29%) | 60% (51% to 69%) | 51% (39% to 62%) | 58% (45% to 71%) | 40% (30% to 52%) | | |
| | 8V | 4% (1% to 10%) | 12% (2% to 24%) | 60% (51% to 69%) | 51% (38% to 61%) | 58% (43% to 71%) | 38% (28% to 48%) | | |
| | 8V-LV | -2% (-9% to 4%) | -2% (-14% to 12%) | 7% (-6% to 19%) | -1% (-18% to 17%) | -7% (-26% to 10%) | -6% (-20% to 9%) | 0 | 1 |
| Mother justifies being beaten for not having sex | LV | 1% (0% to 4%) | 2% (0% to 7%) | 56% (47% to 63%) | 49% (36% to 60%) | 57% (43% to 68%) | 36% (25% to 46%) | | |
| | MV | 3% (0% to 9%) | 7% (0% to 19%) | 58% (49% to 67%) | 57% (45% to 69%) | 55% (43% to 68%) | 40% (29% to 51%) | | |
| | 8V | 2% (0% to 7%) | 2% (0% to 12%) | 58% (49% to 67%) | 51% (38% to 62%) | 51% (38% to 64%) | 36% (25% to 45%) | | |
| | 8V-LV | 1% (-2% to 6%) | 0% (-7% to 10%) | 3% (-9% to 15%) | 1% (-17% to 18%) | -4% (-22% to 13%) | 0% (-14% to 14%) | 0 | 0 |
| Mother justifies being beaten for burning food | LV | 3% (0% to 7%) | 5% (0% to 12%) | 29% (22% to 35%) | 30% (19% to 40%) | 62% (52% to 74%) | 32% (23% to 43%) | | |
| | MV | 6% (1% to 11%) | 10% (2% to 21%) | 30% (22% to 38%) | 32% (22% to 45%) | 51% (36% to 64%) | 28% (17% to 38%) | | |
| | 8V | 3% (0% to 8%) | 2% (0% to 12%) | 26% (19% to 35%) | 31% (19% to 44%) | 51% (38% to 64%) | 28% (17% to 37%) | | |
| | 8V-LV | 0% (-4% to 6%) | -2% (-12% to 7%) | -2% (-13% to 9%) | 1% (-13% to 17%) | -13% (-30% to 6%) | -6% (-18% to 8%) | 0 | 2 |
| Mother justifies being beaten | LV | 30% (21% to 41%) | 33% (19% to 48%) | 70% (63% to 76%) | 52% (40% to 64%) | 87% (78% to 94%) | 53% (43% to 63%) | | |
| | MV | 31% (22% to 42%) | 33% (19% to 50%) | 74% (67% to 82%) | 60% (48% to 71%) | 88% (78% to 96%) | 55% (43% to 66%) | | |
| | 8V | 24% (16% to 34%) | 29% (14% to 45%) | 74% (67% to 82%) | 56% (43% to 68%) | 90% (81% to 97%) | 52% (41% to 62%) | | |
| | 8V-LV | -6% (-20% to 8%) | -5% (-24% to 14%) | 5% (-6% to 15%) | 3% (-13% to 19%) | 3% (-7% to 14%) | -1% (-15% to 14%) | 0 | 0 |
| Mother decides on her own health | LV | 20% (12% to 29%) | 45% (31% to 60%) | 19% (13% to 26%) | 13% (5% to 20%) | 62% (51% to 74%) | 77% (68% to 85%) | | |
| | MV | 28% (19% to 39%) | 33% (19% to 52%) | 22% (15% to 30%) | 16% (8% to 26%) | 52% (39% to 65%) | 74% (64% to 83%) | | |
| | 8V | 29% (19% to 40%) | 45% (29% to 62%) | 23% (15% to 31%) | 18% (9% to 29%) | 51% (36% to 65%) | 85% (75% to 92%) | | |
| | 8V-LV | 9% (-4% to 24%) | 2% (-19% to 21%) | 4% (-4% to 13%) | 5% (-5% to 17%) | -13% (-30% to 4%) | 8% (-3% to 20%) | 0 | 2 |
| Mother decides on large household purchases | LV | 28% (19% to 38%) | 45% (29% to 60%) | 21% (15% to 28%) | 10% (4% to 18%) | 54% (42% to 67%) | 63% (52% to 74%) | | |
| | MV | 28% (19% to 39%) | 26% (12% to 45%) | 23% (15% to 31%) | 16% (8% to 26%) | 43% (30% to 57%) | 69% (57% to 78%) | | |
| | 8V | 31% (22% to 42%) | 38% (21% to 55%) | 20% (13% to 28%) | 16% (8% to 27%) | 48% (33% to 64%) | 72% (61% to 82%) | | |
| | 8V-LV | 3% (-10% to 17%) | -5% (-26% to 14%) | -1% (-9% to 8%) | 5% (-5% to 17%) | -6% (-23% to 10%) | 9% (-5% to 22%) | 0 | 0 |
| Mother decides on visits to family or relatives | LV | 39% (29% to 48%) | 55% (38% to 69%) | 28% (22% to 36%) | 9% (3% to 16%) | 62% (51% to 74%) | 74% (66% to 83%) | 0 | 1 |
| | MV | 33% (23% to 44%) | 29% (14% to 43%) | 38% (29% to 47%) | 14% (6% to 23%) | 51% (38% to 64%) | 74% (63% to 84%) | | |
| | 8V | 40% (29% to 51%) | 38% (21% to 57%) | 31% (23% to 40%) | 14% (6% to 25%) | 55% (41% to 70%) | 80% (70% to 90%) | | |

| | | | | | | | |
|---|---|---|---|---|---|---|---|
| | 8V-LV | 2% (-11% to 16%) | -14% (-36% to 7%) | 2% (-8% to 13%) | 5% (-4% to 16%) | -6% (-23% to 12%) | 7% (-6% to 20%) |

*Data sources: Various DHS [1].*
*Averages were calculated after controlling for the variables in Table 4, through optimal propensity scores matching to obtain a sample of 8V and MV children with similar characteristics to LV children (note LV set is unmodified).*
*† denotes DHS surveys where LV consists of one-year-old children with 0-2 vaccines and MV of 3-7 vaccines.*
*‡ denotes DHS surveys where LV consists of one-year-old children with 0-4 vaccines and MV of 5-7 vaccines.*
** denotes CIs not containing zero. Values correspond to the median of the averages of groups LV, MV, 8V, and 8V-LV differences. Value ranges in parenthesis correspond to the 95% CI obtained by a weighted bootstrap method of 1000 replications.*
